# Changes in out-of-home food purchasing following England’s calorie labelling regulations: a population-level controlled interrupted time series

**DOI:** 10.1101/2025.03.24.25324537

**Authors:** Alexandra Kalbus, Jean Adams, Kerry Ann Brown, Penny Breeze, Alan Brennan, Steven Cummins, Dalya Marks, Cherry Law, Stephen O’Neill, Richard Smith, Oana-Adelina Tanasache, Laura Cornelsen

## Abstract

**Background:** Large out-of-home (OOH) food businesses in England have been required by law to display calorie information on menus since 6^th^ April 2022. This study investigates whether the implementation of this policy was associated with changes in calories purchased OOH.

**Methods:** Controlled interrupted time series analysis was used to estimate changes in calories purchased from all OOH outlets in England (intervention group). Secondary outcomes included purchases from large chain outlets, non-chain outlets and five sub-types of purchases (meals, lower-calorie coffee, higher-calorie coffee, sandwiches, and fish and chip meals). The control series consisted of purchases from non-chain outlets in Scotland and Wales. Itemised OOH food and non-alcoholic drink purchase records, matched to calorie (kcal) content at one point in time, covered 13 weeks pre- and 34 weeks post-intervention (3^rd^ January to 27^th^ November 2022) and were aggregated to population level average weekly kcal purchase estimates. Linear regression, adjusted for season and inflation, was used to model level and trend changes compared to the counterfactual of no mandatory policy. The counterfactual was constructed from the intervention series’ pre-intervention level and the control series’ post-intervention level and trend. Subgroup analyses explored effects by age, sex, socioeconomic status (SES), weight status, and weekday/weekend purchases.

**Results:** Compared to the counterfactual, we found no evidence of a change in overall calories purchased OOH associated with mandatory calorie labelling. There was also no robust evidence of changes in calories purchased OOH for secondary outcomes and by subgroups compared to the counterfactual of no mandatory calorie labelling. Small changes observed in these analyses were sensitive to analytical choices.

**Discussion:** Our findings suggest that mandatory calorie labelling in England did not reduce overall calories purchased from OOH venues. Findings should be interpreted taking into account that the data used did not capture possible changes to menus. Our findings support existing evidence that information provision alone is unlikely to secure significant changes in food purchasing behaviour at population level.

## Introduction

Diet-related diseases are a global public health concern (Meier et al., 2019; The GBD 2015 Obesity Collaborators, 2017). More than 25% of UK adults live with obesity and a further 38% with overweight, with disadvantaged groups disproportionately affected (NHS Digital, 2022). Diets high in energy-dense and nutrient-poor foods are a key contributor to unhealthy weight and poor dietary health. Food prepared away from home contributes on average around 300 kcal per person per day (Mariani et al., 2024) and it tends to be less healthy compared to food prepared at home (Bezerra et al., 2012; Lachat et al., 2012). Meals served in large UK restaurant and fast-food chains have been reported to typically exceed 600 kilocalories (kcal) recommended per serving (Huang, Burgoine, Theis, et al., 2022; Muc et al., 2019; Robinson et al., 2018).

In an effort to improve population diets, some voluntary strategies focussing on out-of-home (OOH) food in England have been proposed over time. The Public Health Responsibility Deal (RD) was launched in 2011 as public-private partnership encouraging businesses to sign up to pledges to work towards improving their practices that could influence population health. These included displaying nutritional information of food served in restaurants and takeaways (Durand et al., 2015). An evaluation of the food-related RD pledges found that actions had limited ‘added value’ as many businesses only committed to pledges they were already compliant with (Durand et al., 2015). However, even where implemented, calorie labelling often did not meet respective recommendations (Robinson et al., 2019). Another voluntary effort followed in 2016 to improve OOH food through reformulation in the Calorie Reduction Programme, but no progress was achieved (UK Government, 2024).

Given the limitations of voluntary measures (Goodman et al., 2018), mandatory calorie labelling in the OOH food sector in England was introduced from 6^th^ April 2022. The calorie labelling regulations apply to all out-of-home food businesses with ≥250 employees that offer prepared food ready for immediate consumption (UK Government, 2021a). These businesses are required to display the calorie content of menu items at the point of choice. The calorie label must correspond to the serving size offered, be displayed in kcal and be accompanied by the statement ‘adults need around 2,000 kcal a day’ (UK Government, 2021a).

Calorie labelling is hypothesised to reduce overall calories consumed, and thereby improve population diet, via two pathways. The first is via individual behaviour change whereby consumers select lower-calorie foods as a result of having access to nutritional information (Agarwal et al., 2021). This requires the consumer to notice this information, gain additional information through the label they would not have had otherwise, and be able and willing to use it to make healthier food choices (Burton & Kees, 2012). The second mechanism is via change in the offer whereby mandating calorie labels to be shown encourages food businesses to change their menus by removing higher-calorie options, offer more lower-calorie options, or reformulate existing dishes to reduce calorie content (Zlatevska et al., 2018).

Most evidence on the effectiveness of calorie labelling to improve diets focusses on the first pathway (individual behaviour change) and originates from the US, where despite a mixed evidence base, an overall significant calorie reduction following calorie labelling has been established by meta-analyses (Agarwal et al., 2021). Zlatevska et al. (2018) conducted a separate meta-analysis for each pathway, based on experimental as well as observational studies, also predominantly from the US. They found that both pathways led to a reduction in calories, with consumers’ food choice associated with a 27 kcal per meal reduction, and changes in the food offered by out-of-home businesses associated with a 15 kcal per meal reduction (Zlatevska et al., 2018).

There is limited knowledge on the calorie labelling regulations’ impact on population health in England. Experimental studies that typically randomise participants into viewing menus with or without calories have found generally lower calories ordered among those who were provided calorie labels (Finlay et al., 2023; Liddiard & Hamshaw, 2024; Tanasache et al., 2025). Colombet et al. (2024) modelled population-level obesity prevalence and cardiovascular disease mortality in England following the implementation of the calorie labelling regulations. Assuming a reduction of 47 kcal per meal suggested by a previous meta-analysis (Crockett et al., 2018; Tanasache et al., 2025), they concluded that as currently implemented, the policy would reduce obesity prevalence by 0.31% over 20 years. Recently published studies conducted in real-world settings examining the effects of the mandatory policy have relied on surveys with limited geographical reach or limited to specific settings such as the workplace: Polden et al. (2024) used pre-post customer intercept design in four areas of England, while Luick et al. (2024) analysed changes in calories purchased in workplace cafeterias using an interrupted time series design. Both found no effects of the policy on consumer behaviour. However, another study found a small overall reduction in calories offered by chain restaurants (Essman, Burgoine, Huang, et al., 2024) suggesting some effectiveness of the mechanism whereby labelling encourages changes through offer. To our best knowledge, there is no research to date considering the effects of the calorie labelling regulations on consumer behaviour at the population level in England.

In this study, we seek to address this knowledge gap by examining changes in OOH food and non-alcoholic drink purchasing following the introduction of mandatory calorie labelling in the OOH food sector in England using a controlled interrupted time series (CITS) design.

## Methods

We adopted a CITS design to estimate the effect of England’s calorie labelling regulations on calories purchased from OOH food and non-alcoholic drinks using transaction-level consumer purchase data from January to November 2022. The CITS design compares observed purchases in England following implementation of calorie labelling on 6^th^ April 2022 (the intervention) with a counterfactual where the policy had not been implemented.

### Data

#### Food and drink expenditure data

We used transaction-level food and drink purchasing data from the Kantar’s Worldpanel Out-of-Home Purchase Panel for the period 3^rd^ January 2022 to 27^th^ November 2022. This contains information on purchases of prepared food and non-alcoholic drinks for consumption away from home or prepared food purchased for at-home consumption (takeaways). Purchases are continuously recorded by a sample of ∼7,500 individuals that are part of Kantar’s Worldpanel Take-home panel of ∼30,000 households across Great Britain who record all take-home purchases. The sample of OOH purchasers are representative in terms of individual characteristics (age group and sex) and region of residence of the population aged 13–79 years in Great Britain.

Panellists record OOH purchases via a mobile phone application. The dataset available for this research included the respective food and drink item’s name, product identifier, and price; the date of purchase; an identifier of the purchase occasion, e.g. where several items were bought together; where it was bought from (store identifier), which includes names of larger retailers and generic categories for smaller businesses, e.g. ‘café’; and who it was bought for, e.g. for the individual themselves or other adults or children.

Kantar provided a dataset of calorie information separately collected from retailer websites. These data were available for one point in time, with most information collected between June and August 2022. For products for which no calorie information was available (68.90% of products), predominantly purchased from small businesses not required to display this information, or products that are on the menus temporarily and are not eligible for labelling regulations, imputed calorie information was provided. These imputed calories were calculated as trimmed means based on similar products in the same outlet or similar types of outlet. For instance, calories for a cheese sandwich sold in a café would have been imputed using calorie information from other sandwiches in the same outlet, if available, or based on cheese sandwiches from other cafés. We linked calorie information and purchase records by product and outlet (97.32% of purchase records), and non-outlet specific products (such as branded bottled drinks) by product identifier only (2.08% of purchase records). For the remaining unmatched purchase records, nutrition information was retrieved from Kantar’s Worldpanel Take-home Panel Data if the products were barcoded, e.g. bottled drinks and packaged snacks (0.07%), looked up manually on businesses’ websites (0.04%) or imputed based on similar products in similar types of outlets (0.50%).

Information on the individuals reporting OOH purchases included their age, sex, occupational socio-economic status (SES), body mass index (BMI) calculated from self-reported height and weight, and region of residence. SES is based on the individual’s occupational social grade following the classification by the National Readership Survey (2018). We operationalised SES as follows: high (AB: “Higher and intermediate managerial, administrative and professional” & C1: “Supervisory, clerical and junior managerial, administrative and professional”) and low (C2: “Skilled manual workers” & DE: “Semi-skilled and unskilled manual workers; and State pensioners, casual and lowest grade workers, unemployed with state benefits only”). SES was unknown for four individuals, who were excluded from the respective subgroup analysis. We used BMI information to determine the individual’s weight status as follows (World Health Organization, 2000): underweight and healthy weight, grouped together due to the low prevalence of underweight in the sample: < 25 kg/m^2^; overweight: 25–29.9 kg/m^2^; obesity: ≥ 30 kg/m^2^. Weight status was unknown for 26% of reporters. Although logistic regression suggests that missingness was not associated with calories purchased, findings of the subgroup analysis by weight status should be interpreted as exploratory. Region of residence was dichotomised into intervention (England) and control group (Scotland & Wales).

#### Outcomes

The primary outcome was population-level mean calories (kcal) purchased from OOH food businesses per person per week. OOH food businesses were defined as any business that may have been subject to the regulations, i.e. offered unpackaged, prepared food and drink ready to be consumed, irrespective of its size. Such businesses include restaurants, cafes, pubs, and takeaways as well as workplace canteens and entertainment venues. Purchases from supermarkets and other retail outlets that were not targeted by the regulations were excluded.

Secondary outcomes included population-level calories per person per week from large chain restaurants and takeaways for which names were included in the dataset (as these are preset in the mobile application where purchases are recorded), referred to hereafter as large chains. These were businesses we could identify by name in the data and were sufficiently large (250+ employees) to be required to show calories. However, they do not include the totality of large OOH food businesses in England, as not all large chains are named in the data. We identified 92 large chains from the data which represented approximately fifth of all the large businesses (250+ employees) in the Accommodation and Food Service sector (Department of Health and Social Care, 2020). We also considered purchases made from OOH food businesses excluding the identified chains, referred to as non-chains. This outcome was chosen to assess the effects of voluntary labelling on smaller businesses from possible substitution from restaurants with calorie labelling to those without (albeit we cannot rule out that some purchases in this group were from large businesses that were not named and therefore not identified as such). Further outcomes included purchases of calories from all meals (pre-defined by Kantar in the purchase data), coffees (distinguishing higher- and lower-calorie options defined as coffees with high milk content such as cappuccino, mocha, latte, and low milk content such as americano, filter coffee, macchiato, cortado, respectively), sandwiches (including baguettes, wraps and other types of bread, but excluding hot dogs and burgers), and fish and chip meals. These outcomes were chosen to reflect a range of products commonly purchased from chains for individual rather than shared consumption, and with sufficient observations available.

#### Analytical dataset

While the policy was mandated only in England, it is highly likely that it had spill-over effects to Scotland and Wales, as some large cross-border food businesses operate across the United Kingdom. Authors’ communication with The Food Foundation and email correspondence with four large restaurant chains confirmed that these businesses implemented calorie labelling uniformly across their UK branches, effectively treating the regulations as UK-wide policy (unpublished author communication). These four businesses contributed 20.2% of OOH purchases in Scotland and Wales. As this violates an identifying assumption for a controlled study – the stable unit treatment value assumption (SUTVA) which states that the control group should be unaffected by the intervention (Kim & Steiner, 2016) – we removed purchases from chains identified in the data from the control series. Purchases from all OOH food outlets in England, the intervention series, are thereby controlled through purchases only made from non-chains (e.g. independent restaurants, cafes and takeaways) in Scotland and Wales. Therefore, for the control series, the outcomes ‘all OOH purchases’ and ‘purchases from non-chains’ were identical.

The study period was restricted to 3^rd^ January to 27^th^ November 2022, corresponding to 13 weeks pre and 34 weeks post intervention, to represent full weeks and to exclude the Christmas period. The intervention was assumed to begin the week including the implementation date (6^th^ April), which started on Monday, 4^th^ April 2022. Data pre-dating January 2022 were not sought due to this period coinciding with measures related to the COVID-19 pandemic still in place restricting interaction with the OOH food sector (UK Government, 2021b).

The analytical dataset included 542,671 purchase records (119,378 pre intervention and 423,293 post intervention) made during 331,966 purchase occasions (74,691 pre and 257,275 post). These were recorded by 6,508 individuals, of which 5,787 (88.9%) resided in England, 462 (7.1%) in Scotland and 259 (4.0%) in Wales. This analytical sample is smaller than the overall OOH panel because we excluded purchases from supermarkets and other retail and removed purchases from chains from the control series.

Purchases of calories were then aggregated to weekly population-level purchases per person using sampling weights provided by Kantar. The weights refer to the population assumed to be purchasing food and drink away from home, consisting of ∼50 million individuals aged 13–79 years in Great Britain and are created by Kantar to ensure representativeness in terms of age, sex, presence of children in the household, and region of residence. They also incorporate weighting specific to underreporting of products and SES. Further details of the weights are Kantar proprietary information, but we confirmed that the weighting process was not changed during the study period and we have thus no reason to expect biases from applying weights (unpublished author communication). We applied weights to create aggregate series of population-level calories purchased OOH per person per week. The final analytical dataset consisted of 47 weekly observations each in the intervention and control series.

#### Covariates

To account for rapid rises in inflation throughout 2022, we included the monthly change rate in Consumer Price Index (CPI) in the UK as a covariate (2024b). We compared model performance when including the overall inflation rate (including non-food expenditures such as other retail and energy prices, which we assumed to reflect the overall strain on household finances) and inflation specific to food, with the former indicating better model fit. To account for seasonality of purchasing, we included indicator variables for spring, summer, autumn, and winter expressed as 3-month periods. We also explored controlling for food-related festivals (Valentine’s, Easter, Queen’s Jubilee, Halloween), but this did not improve the model fit.

### Statistical analysis

We modelled the effect of the calorie labelling regulations on weekly per person calories purchased OOH for each outcome in a CITS design using linear regression. We modelled immediate (level) and trend changes associated with the implementation of calorie labelling (Bernal et al., 2017). Models for all studied outcomes were structured as follows (suppressing coefficients):

Estimated outcome = group + time + intervention + time after intervention + intervention x group + time after intervention x group + season + CPI change + CPI change x group

Group is a binary indicator of whether the observation belongs to the intervention (1) or control series (0). Time (in weeks) was included as linear term, which yielded a better model fit than a quadratic term. The intervention effect was modelled as level change (intervention x group) and trend change (time after intervention x group). Post-intervention trend (time after intervention) was set to 0 at the time of intervention and as 1, 2, …,33 in the following weeks, facilitating interpretation of the intervention’s coefficient as level change. Inflation was allowed to have separate effect in the intervention and control series (CPI x group). Following this model specification, the counterfactual was constructed from a combination of pre-intervention purchasing levels in the intervention group, common pre-intervention trends and post-intervention purchasing trends in the control group. Model performance was assessed by comparing models’ BIC and RMSE.

We observed parallel pre-intervention trends for both an unadjusted and fully adjusted model, indicated also by the non-significant time x group coefficient, which allows pre-intervention time trends to vary between treatment and control group (see model building in Supplementary Material 1: Table S1). We therefore proceeded with the analysis without this interaction term. While there is some lack of consensus in the literature as to whether the inclusion of this term is necessary (Bernal et al., 2018; Bottomley et al., 2019) we follow St.Clair et al. (2016) who argue that if there are parallel trends in the pre-intervention series, this interaction term should not be included to allow greater precision of the effect estimates. We present models including the interaction term allowing varying pre-trends in the robustness checks (see Supplementary Material 4).

Despite the short length of the series, we tested for autocorrelation using the Durbin-Watson test. Where residuals were found to be autocorrelated, we present robust standard errors.

We present level and trend changes for all analyses undertaken. For the main outcome and where intervention effects were observed, we present plots showing observed, predicted and counterfactual values.

#### Subgroup analysis

In further analyses, we explored effect heterogeneity in primary outcome (weekly per-person calories purchased OOH) by available respondent characteristics, namely age (three age bands: <35 years; 35–54 years; 55+ years), sex (male, female), SES (high, low), and weight status (under and healthy weight, overweight, obesity), and whether purchases were made on a weekday or weekend. As these subgroups are generally associated with diet and dietary health (Paddock et al., 2017; Roberts et al., 2018), there is a need to understand whether the impact of calorie labelling varies by these groups. Transaction-level data were separately aggregated to create subgroup datasets of weekly calories purchased in the intervention and control group. The intervention effect was estimated through the same model as described above using the stratified data. For the comparative analysis by weekday or weekend, calories were rescaled to calories purchased per person per day.

#### Robustness checks

We tested the robustness of findings to our methodological choices in various ways. First, we explored how sensitive findings were to our model specification by allowing the pre-intervention trends to vary between treatment and control group. Second, we restricted the purchase records to only those recorded as made for the individual excluding purchases made for other adults or children. This reduced the transaction-level dataset to 57.8% of observations. Third, to assess the influence of outlier observations, we excluded the top 1% and top 10% purchase occasions by calorie content, respectively. Fourth, we used balanced observations by restricting the post series to until 3^rd^ July 2022, leaving 13 weeks each pre and post intervention. Fifth, we explored if the actual date of policy implementation (week 14, starting 4^th^ April) reflected the start of the intervention by moving the start of the modelled intervention backwards by one month, to the week commencing on 7^th^ March 2022. Some outlets may have introduced calorie labels on menus in anticipation of the policy ahead of 6^th^ of April. This is likely to be affected by individual outlet menu change cycles and therefore impossible *a priori* to accurately predict. If the start of the intervention was correctly specified, we would expect to see smaller intervention effects by moving it backwards, particularly in the level change (as the true intervention was still captured in the post-series). Lastly, to assess whether UK-wide CPI sufficiently captured the impact of inflation on OOH food and drink purchases, we included a price index of average price per item purchased OOH (considering items purchased from all OOH outlets) per week and group. This index was not sales-weighted to avoid capturing possible substitution from more expensive to less expensive menu items that might have occurred as a response to inflation.

All data preparation and analysis tasks were carried out in R version 4.4.1, specifically using the packages tidyverse (Wickham et al., 2019), performance (Lüdecke et al., 2021), marginaleffects (Arel-Bundock, 2022), report (Makowski et al., 2023), and rempsyc (Thériault, 2023). Alpha, representing the type 1 error rate, was set as 0.05.

## Results

Table 1 shows descriptive statistics of population-level calories purchased in the intervention and control series. Per-person weekly average calories purchased OOH in the intervention series pre-intervention were 2,348.6 kcal, of which 922.2 kcal were purchased from large chain outlets. Overall, there was a decreasing trend in calories purchased OOH in both intervention and control group, although this was not statistically significant for all outcomes studied. Note that the control series excludes purchases made from large chain outlets and therefore levels are not directly comparable with the intervention series.

**Table 1.**
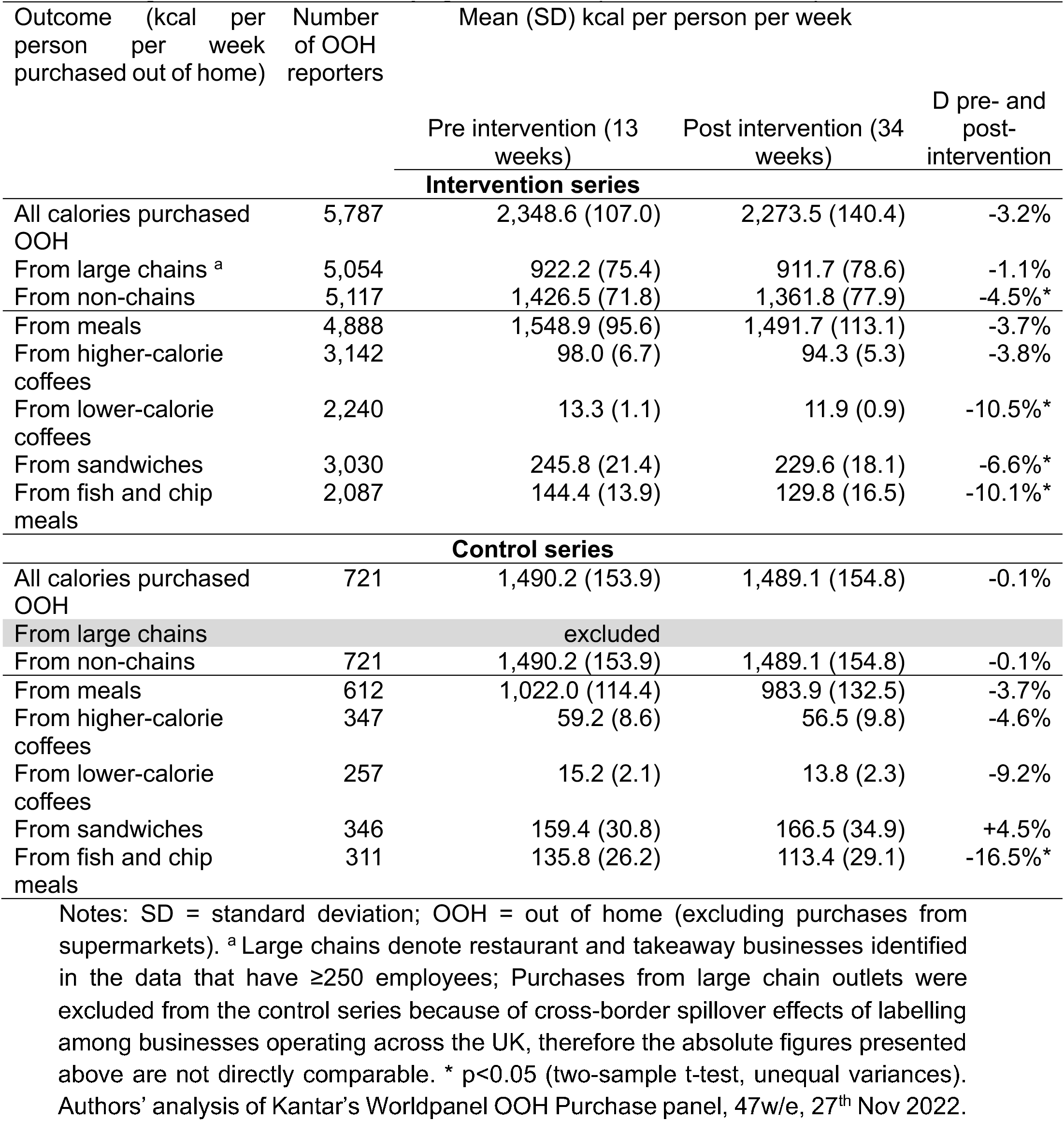
Mean kcal per person per week purchased in intervention and control series before and after labelling introduction and underlying number of reporters in OOH sample.

| Outcome (kcal per person per week purchased out of home) | Number of OOH reporters | Mean (SD) kcal per person per week |  |  |
| --- | --- | --- | --- | --- |
|  |  | Pre intervention (13 weeks) | Post intervention (34 weeks) | D pre- and post-intervention |
| **Intervention series** |  |  |  |  |
| All calories purchased OOH | 5,787 | 2,348.6 (107.0) | 2,273.5 (140.4) | -3.2% |
| From large chains <sup>a</sup> | 5,054 | 922.2 (75.4) | 911.7 (78.6) | -1.1% |
| From non-chains | 5,117 | 1,426.5 (71.8) | 1,361.8 (77.9) | -4.5%* |
| From meals | 4,888 | 1,548.9 (95.6) | 1,491.7 (113.1) | -3.7% |
| From higher-calorie coffees | 3,142 | 98.0 (6.7) | 94.3 (5.3) | -3.8% |
| From lower-calorie coffees | 2,240 | 13.3 (1.1) | 11.9 (0.9) | -10.5%* |
| From sandwiches | 3,030 | 245.8 (21.4) | 229.6 (18.1) | -6.6%* |
| From fish and chip meals | 2,087 | 144.4 (13.9) | 129.8 (16.5) | -10.1%* |
| **Control series** |  |  |  |  |
| All calories purchased OOH | 721 | 1,490.2 (153.9) | 1,489.1 (154.8) | -0.1% |
| From large chains |  | excluded |  |  |
| From non-chains | 721 | 1,490.2 (153.9) | 1,489.1 (154.8) | -0.1% |
| From meals | 612 | 1,022.0 (114.4) | 983.9 (132.5) | -3.7% |
| From higher-calorie coffees | 347 | 59.2 (8.6) | 56.5 (9.8) | -4.6% |
| From lower-calorie coffees | 257 | 15.2 (2.1) | 13.8 (2.3) | -9.2% |
| From sandwiches | 346 | 159.4 (30.8) | 166.5 (34.9) | +4.5% |
| From fish and chip meals | 311 | 135.8 (26.2) | 113.4 (29.1) | -16.5%* |

Notes: SD = standard deviation; OOH = out of home (excluding purchases from supermarkets). <sup>a</sup> Large chains denote restaurant and takeaway businesses identified in the data that have ≥250 employees; Purchases from large chain outlets were
excluded from the control series because of cross-border spillover effects of labelling among businesses operating across the UK, therefore the absolute figures presented above are not directly comparable. \* $p < 0.05$ (two-sample t-test, unequal variances). Authors' analysis of Kantar's Worldpanel OOH Purchase panel, 47w/e, 27<sup>th</sup> Nov 2022.

### Main analysis

Effect estimates (level and trend coefficients) for all outcomes following the final model specification are shown in Table 2. Full model coefficients and graphs are provided in Supplementary Material 2.

**Table 2.** Immediate and trend effects of mandatory calorie labelling at population level.

| Outcome (kcal per person per week out of home) | Level change (95%CI) | P | Trend change (95%CI) | P |
| --- | --- | --- | --- | --- |
| All calories purchased OOH | -95.6 (-471.2 to 280.0) | 0.618 | 5.1 (-5.5 to 15.8) | 0.344 |
| From large chains <sup>a</sup> | -31.2 (-351.8 to 289.4) | 0.849 | **9.2 (0.2 to 18.3)** | **0.046** |
| From non-chains | -155.0 (-466.8 to 156.8) | 0.330 | 4.2 (-4.6 to 13.1) | 0.349 |
| From meals | -78.6 (-390.0 to 232.9) | 0.621 | 8.3 (-0.6 to 17.1) | 0.067 |
| From higher-calorie coffees | -6.3 (-28.0 to 15.3) | 0.566 | 0.4 (-0.2 to 1.1) | 0.161 |
| From lower-calorie coffees | 1.2 (-2.8 to 5.1) | 0.565 | **0.3 (0.2 to 0.4)** | **<0.001** |
| From sandwiches | -31.7 (-106.4 to 43.0) | 0.405 | -2.0 (-4.1 to 0.2) | 0.070 |
| From fish and chip meals | 13.2 (-53.3 to 79.7) | 0.697 | 0.1 (-1.8 to 2.0) | 0.893 |

95%CI = 95% confidence interval; OOH = out of home (excluding purchases from supermarkets). <sup>a</sup> Large chains denote restaurant and takeaway businesses identified in the data that have $\geq 250$ employees. Authors' analysis of Kantar's Worldpanel OOH Purchase panel, 47w/e, 27<sup>th</sup> Nov 2022.

All of the following changes are with respect to the counterfactual where mandatory calorie labelling had not been implemented. We observed no evidence of changes in calories purchased OOH. Observed, predicted and counterfactual values for calories purchased OOH are shown in Figure 1. There was some evidence of an increasing trend in calories purchased from chains by 9 kcal per person per week (95%CI 0.15 to 18.32) following the implementation of calorie labelling (see Figure 2). Further, there was strong evidence of an increasing trend in calories purchased from lower-calorie coffees purchased following the intervention (0.3 kcal per person per week, 95%CI 0.19 to 0.41) (see Figure 3). We did not find statistically significant changes at conventional levels in calories purchased OOH in any of the remaining outcomes either as immediate (level) or trend changes associated with mandatory calorie labelling.

**Figure 1.**
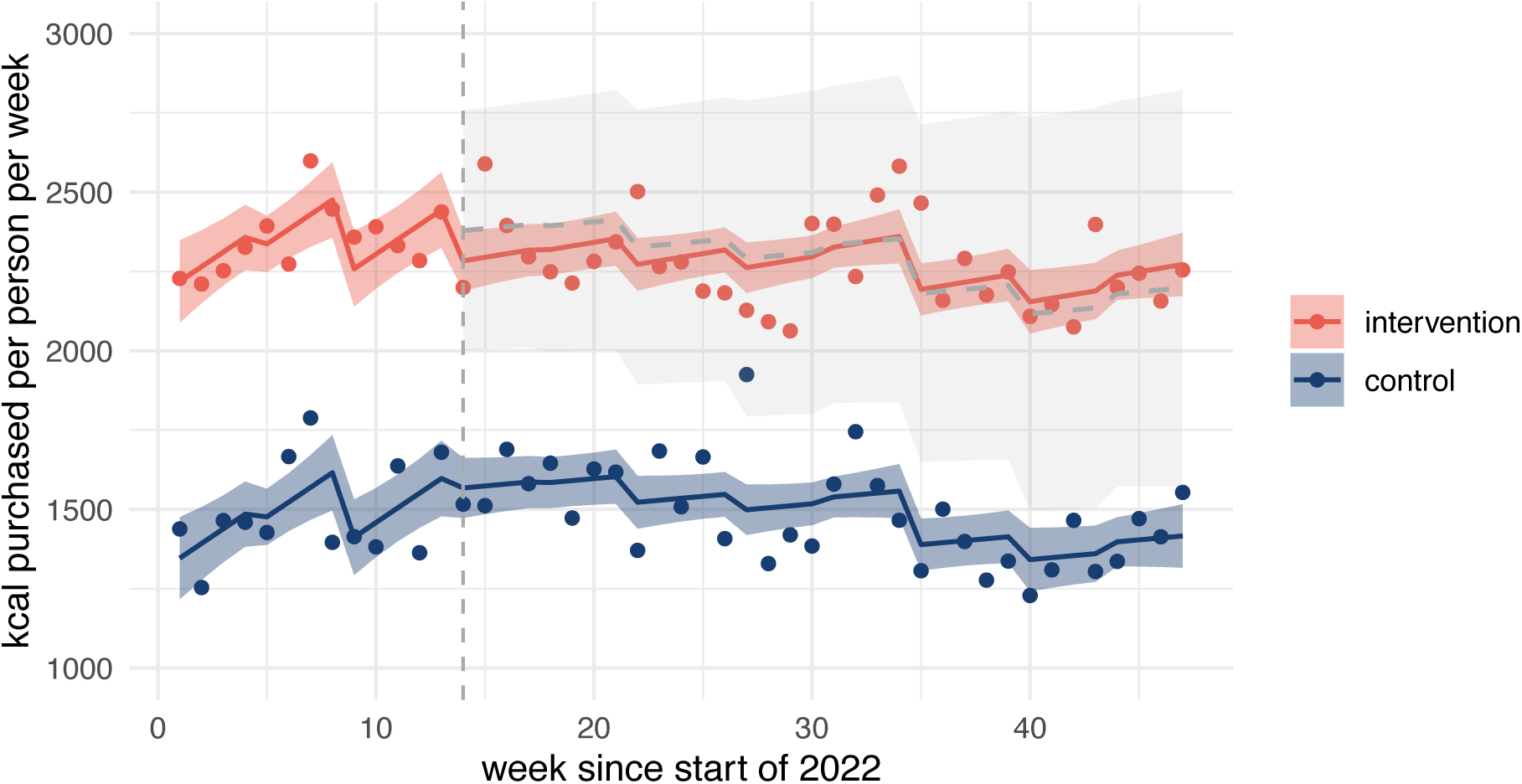
Population-level observed (points) and predicted (solid lines) calories purchased out of home (OOH) with counterfactual (grey dashed line) and 95% confidence intervals (ribbons). Implementation of mandatory calorie labelling = week 14. The intervention series includes all OOH purchases in England. The control series is constructed from purchases made in Scotland and Wales and excludes purchases from large chains. Large chains denote restaurant and takeaway businesses identified in the data that have ≥250 employees. Authors’ analysis of Kantar’s Worldpanel OOH Purchase panel, 47w/e, 27^th^ Nov 2022.

**Figure 2.**
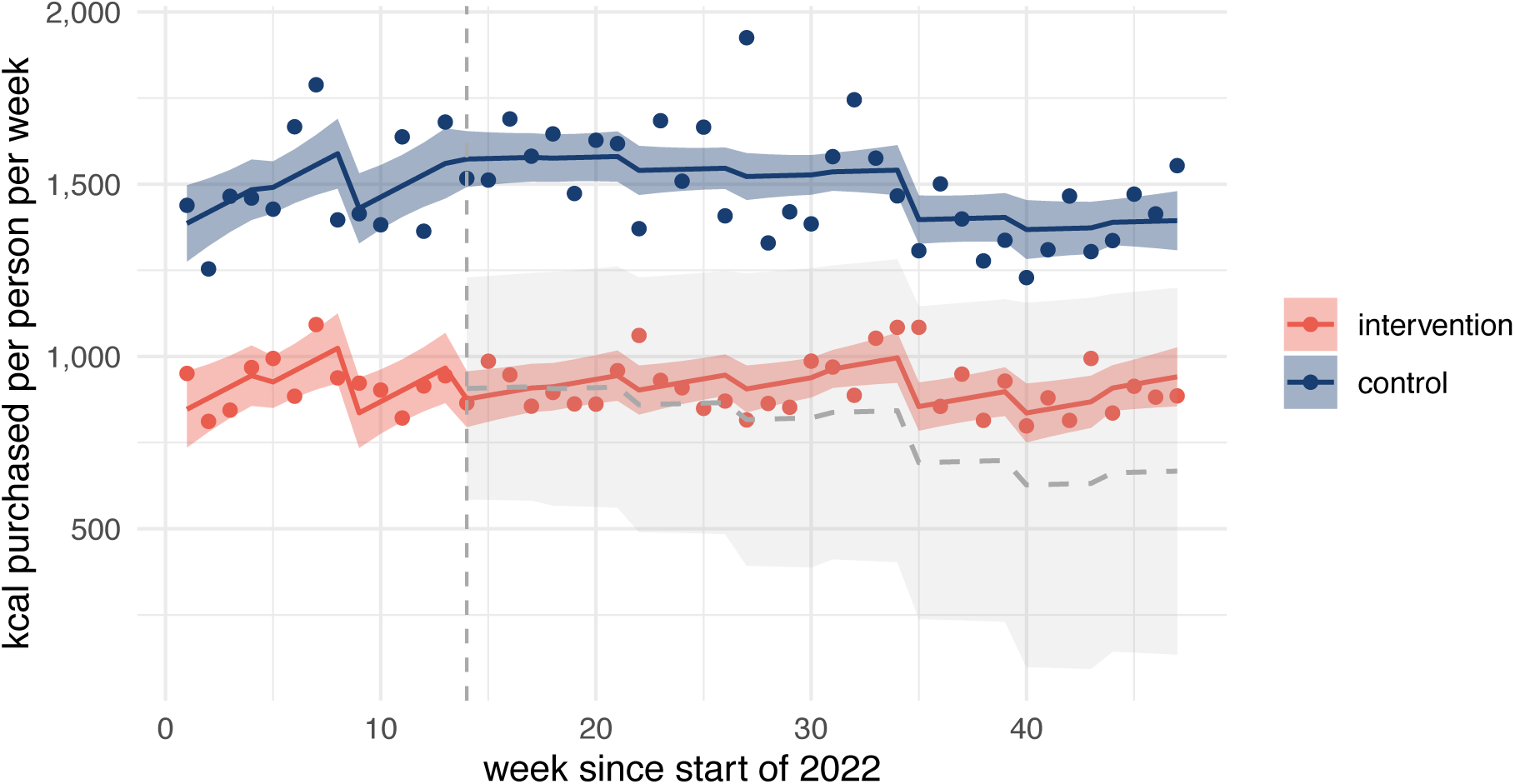
Population-level observed (points) and predicted (solid lines) calories per person purchased from large out-of-home (OOH) chains with counterfactual (grey dashed line) and 95% confidence intervals (ribbons). Implementation of mandatory calorie labelling = week 14. Large chains denote restaurant and takeaway businesses identified in the data that have ≥250 employees. The intervention series includes purchases made from large OOH chains in England. The control series is constructed from purchases made in Scotland and Wales and excludes purchases from large chains. Authors’ analysis of Kantar’s Worldpanel OOH Purchase panel, 47w/e, 27^th^ Nov 2022.

**Figure 3.**
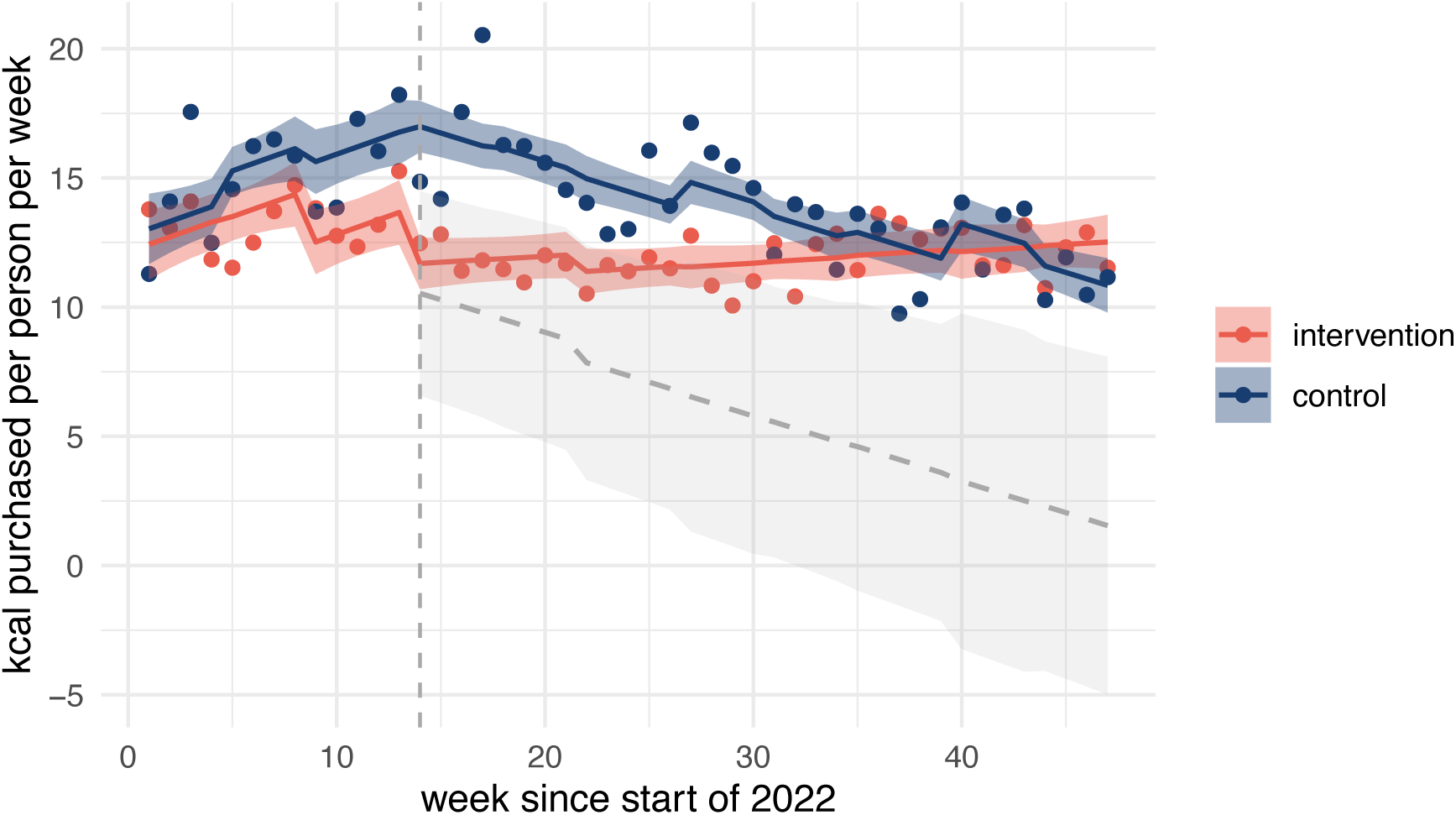
Population-level observed (points) and predicted (solid lines) calories per person purchased from lower-calorie coffees with counterfactual (grey dashed line) 95% confidence intervals (ribbons). Implementation of mandatory calorie labelling = week 14. The intervention series includes purchases in England. The control series is constructed from purchases made in Scotland and Wales and excludes purchases from large chains. Large chains denote restaurant and takeaway businesses identified in the data that have ≥250 employees. Authors’ analysis of Kantar’s Worldpanel OOH Purchase panel, 47w/e, 27^th^ Nov 2022.

#### Subgroup analysis

A descriptive summary of calories purchased OOH by subgroup is provided in Supplementary Material 3. We observed no evidence of effects of mandatory calorie labelling on calories purchased OOH in the analysed subgroups by age, sex, SES, and whether it was a weekday or weekend. However, we found some evidence for an increasing trend in calories purchased OOH following the intervention of 14 kcal per person per week (95%CI 0.4 to 28.3) among people living with overweight (see Table 3 and Figure 4).

**Figure 4.**
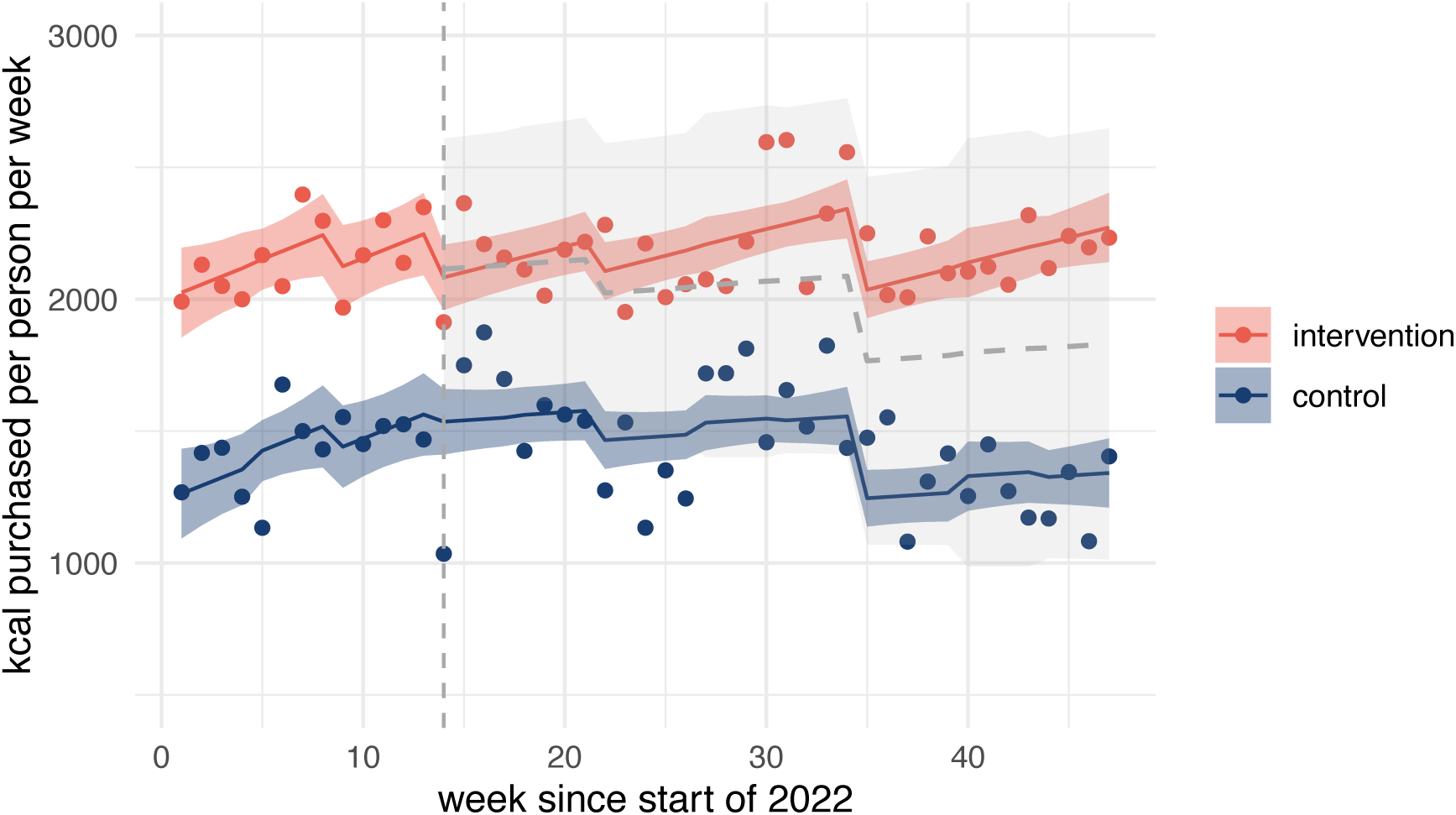
Population-level observed (points) and predicted (solid lines) kcal per person purchased out of home (OOH) by individuals with overweight with counterfactual (grey dashed line) and 95% confidence intervals (ribbons). Implementation of mandatory calorie labelling = week 14. The intervention series includes all OOH purchases in England. The control series is constructed from purchases made in Scotland and Wales and excludes purchases from large chains. Large chains denote restaurant and takeaway businesses identified in the data that have ≥250 employees. Authors’ analysis of Kantar’s Worldpanel OOH Purchase panel, 47w/e, 27^th^ Nov 2022.

**Table 3.**
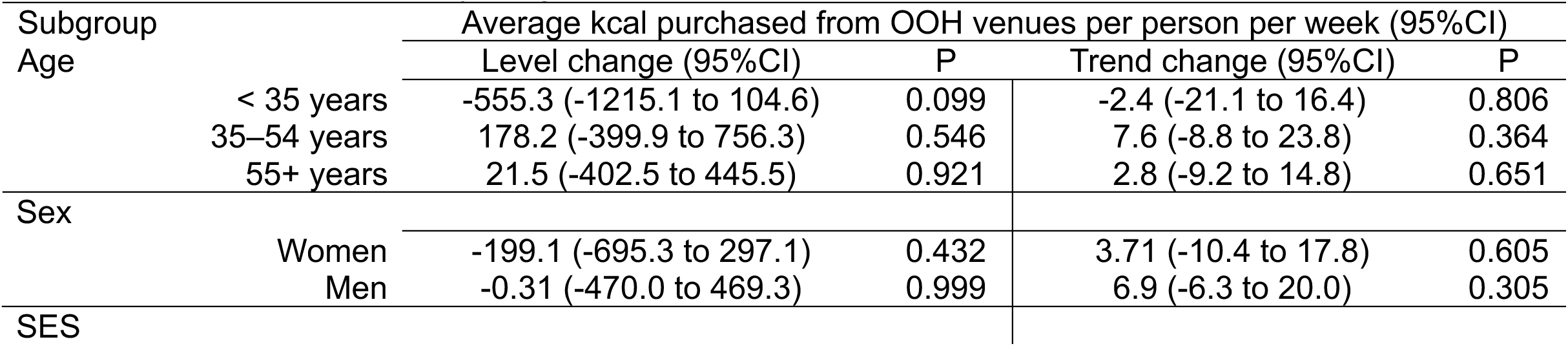
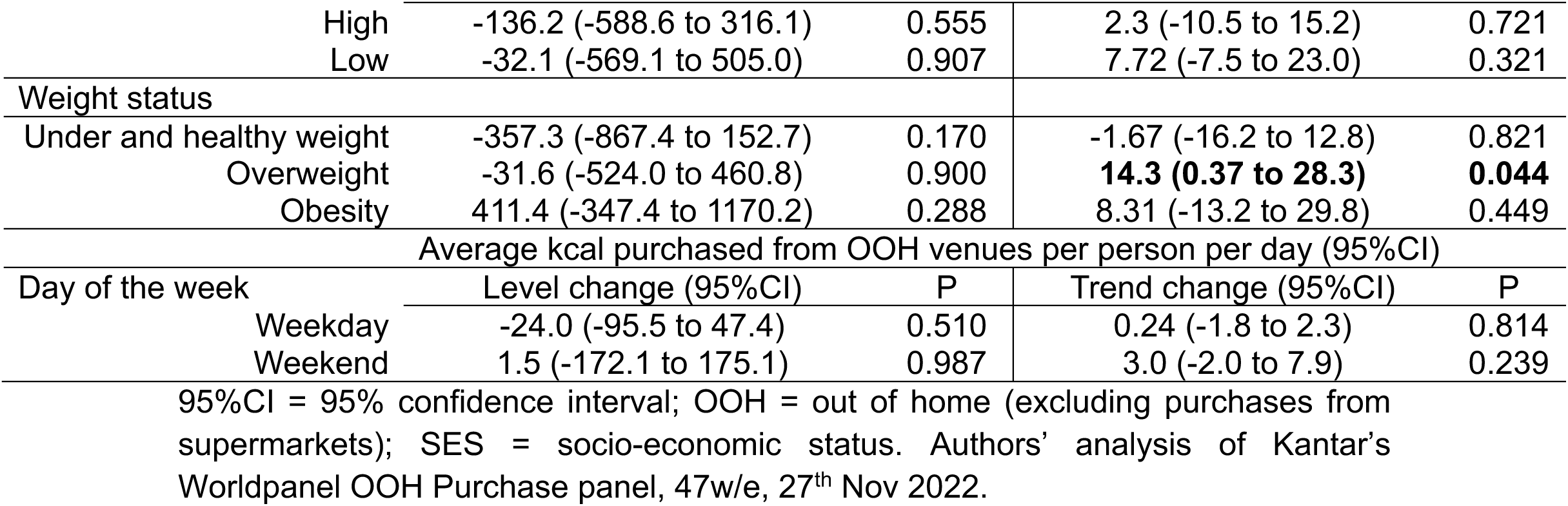
Immediate and trend effects of mandatory calorie labelling on calories purchased from OOH venues per person and week by subgroup.

#### Robustness checks

To understand the robustness of our findings to analytical choices made, we undertook several robustness checks. First, we included an interaction term that allows pre-intervention trends to vary by group (Supplementary Material 4, Table S3). Point estimates were broadly in line with our main analysis, while confidence intervals, as expected, were wider, leading to no statistically significant intervention effects indicated through this analysis.

Second, we considered only purchases made for the individual themselves (Supplementary Material 4, Table S4). Effects for the main outcome (calories purchased OOH) were similar, as was the positive trend change in calories purchased from lower-calorie coffees. However, the trend change in calories purchased from large chains previously observed was reduced by half and was no longer statistically significant. In addition, this analysis indicated an increase in calories from OOH meals (3.7 kcal person/week, 95%CI 0.01 to 7.5) and higher-calorie coffees (0.47 kcal person/week, 95%CI 0.06 to 0.89) but a decrease of calories purchased from sandwiches (-2.5 kcal person/week, 95%CI -4.3 to -0.64).

Third, we excluded the top 1% and 10% of purchase occasions by calorie content. Excluding the top 1% of occasions yielded results similar to those observed in the main analysis (Supplementary Material 4, Table S5). The only difference was that we did not observe the trend change in calories purchased from large chains. Excluding the top 10% of purchase occasions by calorie content also led to similar results for the main outcome and lower-calorie coffee (Supplementary Material 4, Table S6), while the increasing trend change for calories purchased from large chains was not observed. However, this analysis indicated an immediate (level) reduction of 97 kcal per person (95%CI -188.8 to -5.2) from OOH meals overall, and a negative trend change in calories purchased from fish and chip meals (-1.3 (95%CI -2.0 to -0.6).

Fourth, using balanced observations, i.e. restricting the post-intervention period to 13 weeks until Sunday, 3^rd^ July 2022, also resulted in different effect sizes to the main analysis (Supplementary Material 4, Table S7). However, none of the coefficients were significant at conventional levels with confidence intervals crossing zero for all estimates.

Fifth, when moving the intervention start date earlier to 7^th^ March 2022, no intervention effects were observed except the positive trend change in calories from lower-calorie coffee, with a marginally smaller coefficient estimate compared to the main analysis (Supplementary Material 4, Table S8).

Finally, a further adjustment for prices by including the average price per 100 kcal of items purchased OOH (without sales-weighting) led to a moderately better model fit for half of the outcomes studied, including the main outcome. However, the effect estimates were very similar to those from the main analysis (Supplementary Material 4, Table S9).

## Discussion

### Summary of findings

This study aimed to evaluate the impact of implementing mandatory calorie labelling in large food businesses in England on population-level OOH food and drink purchasing. Using transaction-level purchase data in a CITS design, we found limited robust evidence of changes in consumer purchasing following implementation of the calorie labelling regulations. While there was no evidence for an intervention effect on overall calories purchased OOH, we observed increases in calories purchased following the policy from large chains as well as from lower-energy coffee compared to the counterfactual. However, these findings were sensitive to the analytical specifications tested in robustness checks and should thus be interpreted with caution. Exploratory subgroup analyses were mostly in agreement with the main finding of no changes in overall calories purchased OOH, except for an increasing trend in purchases among individuals living with overweight.

### Comparison with other studies

The evidence on the effectiveness of calorie labelling on menus remains mixed. A previous Cochrane systematic review and meta-analysis of the effectiveness of calorie labelling reported a reduction of 47 kcal per meal as a result of calorie labelling (Crockett et al., 2018). This estimate has recently been updated to a reduction of 1.8% in calories selected, corresponding to 11 kcal of a typical meal (Clarke et al., 2025). However, the studies that informed this estimate were predominantly conducted in laboratory settings in the US, included workplace settings and also included visual calorie labels as well as physical activity calorie equivalent (PACE) labels. In contrast, an earlier review of high-quality real-world studies with the majority employing text-based labels akin to those mandated by the English calorie labelling regulations, found no change in calories selected (Kiszko et al., 2014). Findings from these reviews may not be directly applicable to England because of differences in food environments and how individuals interact with them as well as differences in how calorie labels are presented in terms of text/visual label and business compliance.

Evidence on the effectiveness of the calorie labelling regulations in England is emerging. A natural experiment assessing calories per item purchased in workplace canteens reported no change in calories purchased, but found small reductions in the mean calorie content of menu options at each menu change (Luick et al., 2024). Another study using restaurant customer intercept surveys in four local areas before and after implementation of mandatory calorie labelling reported an increase in noticing calorie labels but found no changes in calories purchased or consumed (Polden, Jones, Essman, Adams, Bishop, Burgoine, Sharp, et al., 2024). Our study, finding no evidence of an impact of mandatory calorie labelling on calories purchased OOH, aligns with this emerging evidence base in England.

We observed only limited evidence of differential effects of the calorie labelling regulations across subgroups, with the only observed effect an increase in calories purchased OOH over time among individuals with overweight. Our subgroup findings have to be interpreted with caution as Kantar sample weights are not constructed to reflect this subgroup split and information on weight status was missing for 26% of underlying reporters. Nevertheless, our findings for age and SES are broadly in line with previous research which has not found effect variation by sociodemographic characteristics. A pooled analysis of 12 randomised controlled trials, of which 5 were UK-based, showed no effect modification by participants’ demographics or other characteristics, including age and socioeconomic status (Robinson et al., 2023). Experimental studies on the effects of calorie labelling on takeaway purchasing in England found no differential impacts by individual characteristics (Finlay et al., 2023; Tanasache et al., 2025). A real-world customer intercept study in England also did not find evidence of varying effects of calorie labelling by age, gender, ethnicity or SES (Polden, Jones, Essman, Adams, Bishop, Burgoine, Sharp, et al., 2024). This may be because differential impacts of calorie labelling may be more related to individual value orientation and less to sociodemographic characteristics and more to (Berry et al., 2019; Brissette et al., 2013).

### Interpretation of findings

The present study adds to the emerging evidence base finding no impact of mandatory calorie labelling on menus alone on consumer behaviour change in England. We did observe some evidence, paradoxically, for an increasing trend in calories purchased from large OOH chains following the implementation of mandatory calorie labelling. This finding, however, needs to be interpreted with caution as it was not robust to analytical choices. While the increasing trend in lower-calorie coffees is seemingly in line with our hypothesis of substitution away from higher-calorie coffees, we did not observe a decrease in purchases of the latter. This finding was further not robust to the model specification of varying pre-intervention trends by group. We also observed an increasing trend in calories purchased OOH among individuals with overweight. However, this is supported only by moderate evidence (p=0.044), constructed from underlying data with a high rate of missing values (26%) and belongs to the set of exploratory subgroup analyses. Further research should specifically examine the relationship between mandatory calorie labelling, behaviour change and weight status in England.

Though not a primary objective of the study, we observed decreases in calories purchased OOH in both intervention and control group following on from a steep increase in purchasing at the beginning of 2022. This initial increase may be explained by the start of the year marked by exiting various COVID-19 pandemic-related restrictions affecting social and public life as well as individuals’ engagement with the OOH food sector (UK Government, 2021b). The following decrease may be a stabilising of behaviour and/or a response to inflation, which spiralled in spring 2022 (Office for National Statistics, 2022).

We may have underestimated the effect of mandatory calorie labelling in our study due to study design elements. Specifically, the policy was not a sharp implementation in the sense that some businesses already provided calorie information, including in the control group (Food Standards Scotland, 2021). Prior to the policy, in August– December 2021, 21% of such businesses provided calorie labels, albeit inconsistently (Polden et al., 2023). As such, we estimated the effect of mandatory calorie labelling compared to no and voluntary labelling. We further estimated an intention-to-treat effect, as not every individual in the intervention group was exposed to calorie labelling due to business eligibility, compliance or individuals noticing calorie labels, as discussed in more detail below.

Notwithstanding these considerations, our study’s findings are in line with the emerging literature on the calorie labelling regulations’ impact in England, as outlined above. A reason for the calorie labelling regulations’ lack of impact may be its reliance on behaviour change. The high level of individual agency required to benefit from calorie labelling, namely health numeracy skills to understand and utilise calorie labels as a guide to healthier eating as well as a drive and willingness to change behaviour, may explain the policy’s limited effectiveness (Adams et al., 2016). If an OOH meal is seen as a treat (Keeble et al., 2022), it is unlikely that calorie content is an important driver of choice. Yet, exemplified by the RD a decade ago, businesses favoured calorie labelling and other interventions focussed on information provision, awareness raising and communication with consumers (Knai et al., 2015). Such interventions are known to be less effective than structural changes involving pricing, reformulation and advertising restrictions (Knai et al., 2015).

Another explanation, particularly pertaining to English context, may be the extent of exposure to calorie labelling dependent on 1) how many businesses are eligible for calorie labelling; 2) business compliance with the regulations; and 3) customers noticing labels. In 2018, businesses with ≥250 employees represented 17% of total outlets in the Accommodation and Food Service sector but nearly half (49%) of the sector’s turnover (Department of Health and Social Care, 2020). Of those outlets under the legislation scope, a fifth did not provide calorie information at all and only 15% met all guidelines of showing calories, with non-compliance particularly in relation to presenting information clearly and prominently (Polden, Jones, Essman, Adams, Bishop, Burgoine, Donohue, et al., 2024). Additionally, even if calorie labels are present, these go widely unnoticed by consumers. Polden et al. (2024) recorded only 30% of restaurant and takeaway customers noticing calorie labels after implementation of calorie labelling. Consequently, even if calorie labelling were effective in reducing calories purchased, it is likely that its effect would have been diluted as far as to not be detectable at population level.

The limited evidence of the calorie labelling regulations’ effectiveness on reducing overall calorie intake is juxtaposed by the opposition to the policy by people with lived experience of eating disorders due to concerns of relapsing into more severe periods of disordered eating (Public Health Scotland, 2024). Qualitative research among people with experience of eating disorders in England also suggests that calorie labelling adversely impacts eating disorders and/or recovery (Frances et al., 2024). In this context, the risks posed to people with eating disorders need to be weighed against the efficacy of calorie labelling identified.

### Limitations

While CITS is a strong design for inferring causality (Chan et al., 2022), the following limitations pertain to our study. First, the key identifying assumption of no interference, or SUTVA (Kim & Steiner, 2016), may have been affected. Although calorie labelling is only mandatory in England, several large cross-border chains have introduced calorie labels uniformly across Great Britain. We reduced the bias from such spill-over effects by removing purchases from identified large businesses from the control series. However, we cannot rule out that some outlets showing calories remained in the control series as not all purchases could be identified by business name. Second, the common shock assumption, which requires idiosyncratic shocks in the post-intervention period to be similar in treatment and control group (Nianogo et al., 2023), may have been violated as the calorie labelling regulations’ implementation coincided with a period of high inflation (Office for National Statistics, 2024a) which could have impacted the frequency, level and types of OOH purchases differentially. As only UK-wide inflation data are available, we had to assume that inflation rates were the same in intervention (England) and control series (Scotland and Wales). To mitigate this, we allowed the effects of inflation on calories purchased to vary by group. We included a further, group-specific price index as a robustness check and found that it did not change our findings. Finally, we estimated changes in calories purchased due to changes in behaviour only. As calorie information was available only for one point in time, our analyses did not capture reductions in purchased calories due to possible concurrent changes in calories offered by food businesses (Essman, Burgoine, Huang, et al., 2024).

## Conclusion

This study used a natural experiment design to assess the impact of the calorie labelling regulations in England on calories purchased from out-of-home food venues. No robust evidence of changes in purchasing of out-of-home foods and non-alcoholic drinks related to the policy were observed, which is in line with existing evidence that calorie labelling alone is unlikely to secure significant changes in food purchasing behaviour. Further research may ascertain if the policy led to a reduction in calories offered by restaurants and thereby led to changes in overall calorie consumption.

## Data Availability

Kantar OOH Service Data used in this study cannot be shared but can be obtained from Kantar (https://www.kantarworldpanel.com/). Data were available for 1st January to 31st December 2022. The analyses used data from 3rd January to 27th November 2022. Analyses and interpretation were conducted independently of Kantar. Kantar can neither independently verify the findings nor endorse the views or findings of this study. While data are sourced from Kantar, they are augmented by data from additional sources.

## DATA STATEMENT

Kantar OOH Service Data used in this study cannot be shared but can be obtained from Kantar (https://www.kantarworldpanel.com/). Data were available for 1^st^ January to 31^st^ December 2022. The analyses used data from 3^rd^ January to 27^th^ November 2022. Analyses and interpretation were conducted independently of Kantar. Kantar can neither independently verify the findings nor endorse the views or findings of this study. While data are sourced from Kantar, they are augmented by data from additional sources.

## FUNDING

This study is funded by the National Institute for Health and Care Research (NIHR) School for Public Health Research (SPHR) (Grant Reference Number NIHR 204000). JA was supported by the Medical Research Council (Grant Reference Number MC UU 00006/7). The views expressed are those of the authors and not necessarily those of the NIHR or the Department of Health and Social Care. Kantar OOH Service Data were partially acquired via project grant (NIHR133887) from the NIHR.

## ETHICAL APPROVAL

Ethical approval was not required as the data were obtained in anonymised format (see https://www.kantarworldpanel.com/).

## CONFLICTS OF INTEREST

The authors declare no conflict of interest.

## ACKNOWLEDGEMENTS

The authors wish to thank colleagues from The Food Foundation, particularly Chloe MacKean, for liaising with large OOH food businesses over implementation of calorie labelling within their organisation.

## ABBREVIATIONS

95%CI: 95% confidence interval
BMI: Body Mass Index
CITS: controlled interrupted time series
CPI: Consumer Price Index
OOH: out-of-home
RD: Public Health Responsibility Deal
SES: Socioeconomic status
SUTVA: stable unit treatment value assumption

## Notes

### Competing Interest Statement

The authors have declared no competing interest.

