## Supplementary Material 1 for "Changes in out-of-home food purchasing following England’s calorie labelling regulations: a population-level controlled interrupted time series"

### Model Building

**Table S1**. Model coefficients (95% confidence interval) for different specifications predicting all calories purchased OOH

| | **Unadjusted model with varying pre-intervention trends** | | **Fully adjusted model with varying pre-intervention trends** | | **Final model – fully adjusted without varying pre-intervention trends** | |
|---|---|---|---|---|---|---|
| | Coefficient (95% confidence interval) | P | Coefficient (95% confidence interval) | P | Coefficient (95% confidence interval) | P |
| Intervention x group (level change) | -143.4 (-402.0 to 115.1) | 0.273 | -67.8 (-477.1 to 341.5) | 0.743 | -95.6 (-476.9 to 285.7) | 0.619 |
| Time after intervention x group (trend change) | 3.6 (-25.5 to 32.6) | 0.808 | -1.2 (-35.9 to 33.4) | 0.943 | 5.1 (-5.7 to 16.0) | 0.347 |
| Group | 855.7 (655.6 to 1,055.8) | **<0.001** | 1,133.3 (-79.5 to 2,346.0) | 0.067 | 968.3 (108.7 to 1,827.9) | **0.028** |
| Time | 11.9 (-8.2 to 31.9) | 0.242 | 42.0 (6.1 to 77.9) | **0.022** | 46.3 (18.2 to 74.4) | **0.002** |
| Time x group | 0.45 (-27.8 to 28.7) | 0.975 | 8.6 (-35.8 to 53.0) | 0.701 | | |
| Intervention | 39.9 (-143.0 to 222.6) | 0.666 | 66.3 (-235.0 to 367.5) | 0.663 | 80.2 (-210.8 to 371.1) | 0.585 |
| Time after intervention | -19.4 (-39.9 to 1.12) | 0.065 | -36.9 (-69.4 to -4.5) | **0.026** | -40.1 (-67.9 to -12.3) | **0.005** |
| Season (spring) | | | -187.1 (-388.5 to 14.3) | 0.068 | -187.1 (-387.4 to 13.2) | 0.067 |
| Season (summer) | | | -250.4 (-490.5 to -10.3) | **0.041** | -250.4 (-489.2 to -11.6) | **0.040** |
| Season (autumn) | | | -409.7 (-717.4 to -101.9) | **0.010** | -409.7 (-715.7 to -103.6) | **0.009** |
| CPI change | | | -60.9 (-222.3 to 100.6) | 0.456 | -78.1 (-212.2 to 56.10) | 0.250 |
| CPI change x group | | | -51.9 (-275.8 to 172.0) | 0.646 | -17.5 (-153.1 to 118.2) | 0.799 |
| Constant | 1,419.1 (1277.7 to 1560.6) | **<0.001** | 1,693.2 (807.0 to 2579.4) | **<0.001** | 1,775.7 (1,002.5 to 2,549.0) | **<0.001** |
| Adjusted $R^2$ | 0.90 | | 0.91 | | 0.91 | |
| BIC | 1,222.6 | | 1,230.9 | | 1,226.5 | |

CPI = consumer price index; OOH = out of home. Authors' analysis of Kantar's Worldpanel OOH Purchase panel, 47w/e, 27th Nov 2022.
