## Supplementary Material 2 for "Changes in out-of-home food purchasing following England’s calorie labelling regulations: a population-level controlled interrupted time series"

### Full population analysis

This is the output (model coefficients and plots) from the main analysis of changes in out-of-home (OOH) purchasing following the introduction of mandatory calorie labelling in the OOH sector at the population level.

This is the authors' analysis of Kantar's Worldpanel OOH Purchase panel, 47w/e, 27th Nov 2022. Purchases were aggregated to the population level using gross-up weights provided with the purchase records. They were then divided by the sample weights to calculate kcal per person per week.

Models are set up as follows (suppressing coefficients): predicted outcome = group + Time + Intervention + Time after Intervention + Intervention x group + Time after Intervention x group + season + CPI change + CPI change x group

Purchases from large OOH chains were excluded from the control series because of cross-border spillover effects of labelling among businesses operating across the UK. Large chains denote restaurant and takeaway businesses identified in the data that have ≥250 employees.

### Calories purchased out of home

These venues include all that could potentially be affected by labelling, i.e. restaurants and takeaways as well as leisure venues such as cinemas and hotels. Purchases from supermarkets were excluded. This outcome includes purchases from venues that are required to show calories and those that are not.

This is the primary outcome in this study.

*All calories (kcal) per person per week purchased OOH*

| Parameter | Fit | *b* | 95% CI (*b*) | *t* | *df* | *p* | *b** | 95% CI (*b**) |
|---|---|---|---|---|---|---|---|---|
| (Intercept) | | 1,775.72 | [1002.49, 2548.95] | 4.57 | 82 | < .001*** | 0.55 | [0.09, 1.00] |
| reg | | 968.27 | [108.69, 1827.85] | 2.24 | 82 | .028* | 0.95 | [0.52, 1.38] |
| Time | | 46.28 | [18.21, 74.35] | 3.28 | 82 | .002** | 1.47 | [0.58, 2.36] |
| Intervention | | 80.18 | [-210.80, 371.16] | 0.55 | 82 | .585 | 0.08 | [-0.22, 0.39] |
| Timeafter | | -40.10 | [-67.92, -12.27] | -2.87 | 82 | .005** | -1.05 | [-1.77, -0.32] |
| Intervention reg | | -95.60 | [-476.85, 285.66] | -0.50 | 82 | .619 | -0.11 | [-0.54, 0.32] |
| Timeafter Intervention reg | | 5.14 | [-5.67, 15.95] | 0.95 | 82 | .347 | 0.12 | [-0.13, 0.37] |
| season [spring] | | -187.09 | [-387.41, 13.22] | -1.86 | 82 | .067 | -0.44 | [-0.90, 0.03] |
| | | | [-489.18, | | | | | [-1.14, |

| | | | | | | | | |
|---|---|---|---|---|---|---|---|---|
| season [summer] | | -250.39 | -11.61] | -2.09 | 82 | .040* | -0.58 | -0.03] |
| season [autumn] | | -409.65 | [-715.70, -103.59] | -2.66 | 82 | .009** | -0.95 | [-1.67, -0.24] |
| CPI change | | -78.06 | [-212.22, 56.10] | -1.16 | 82 | .250 | -0.36 | [-0.83, 0.12] |
| reg × CPI change | | -17.46 | [-153.13, 118.21] | -0.26 | 82 | .799 | -0.04 | [-0.32, 0.24] |
| AIC | 1,193.46 | | | | | | | |
| AICc | 1,198.01 | | | | | | | |
| BIC | 1,226.53 | | | | | | | |
| R2 | 0.92 | | | | | | | |
| R2 (adj.) | 0.91 | | | | | | | |
| Sigma | 128.93 | | | | | | | |

Authors' analysis of Kantar's Worldpanel OOH Purchase panel, 47w/e, 27th Nov 2022.

Autocorrelation:

```
## OK: Residuals appear to be independent and not autocorrelated (p = 0.868).
```

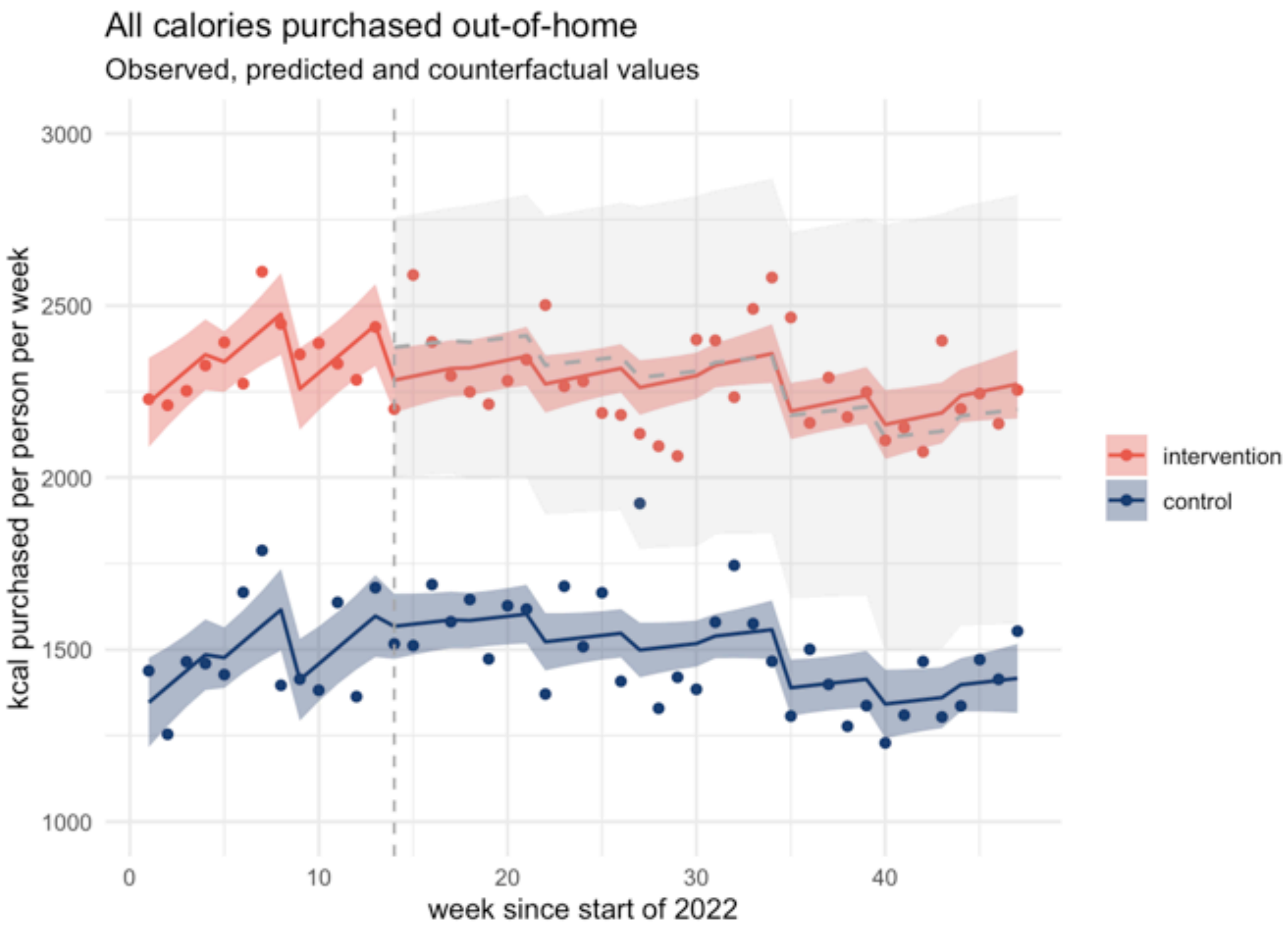


Population-level observed (points) and predicted (solid lines) calories purchased out of home (OOH) with counterfactual (grey dashed line) and 95% confidence intervals (ribbons). Implementation of mandatory calorie labelling = week 14. The intervention series includes all OOH purchases in England. The control series is constructed from purchases made in Scotland and Wales and excludes purchases from large chains. Large chains denote restaurant and takeaway businesses identified in the data that have ≥250 employees. Authors' analysis of Kantar's Worldpanel OOH Purchase panel, 47w/e, 27th Nov 2022.

### Purchases from chains

This outcome includes purchases made from food outlets which can be identified in the data as big chain restaurants and takeaways required to show calorie information. Note that the control series includes purchases from all outlets except known chains (therefore the same control series as shown in all OOH purchases above).

*Purchases (kcal) per person per week from large OOH chains*

| Parameter | Fit | *b* | 95% CI (*b*) | *t* | *df* | *p* | *b** | 95% CI (*b**) |
|---|---|---|---|---|---|---|---|---|
| (Intercept) | | 1,587.68 | [927.69, 2247.66] | 4.79 | 82 | < .001*** | 0.61 | [0.07, 1.14] |
| reg | | -341.24 | [-1074.93, 392.46] | -0.93 | 82 | .358 | -1.06 | [-1.57, -0.56] |
| Time | | 32.55 | [8.59, 56.51] | 2.70 | 82 | .008** | 1.42 | [0.37, 2.46] |
| Intervention | | 53.50 | [-194.86, 301.86] | 0.43 | 82 | .669 | 0.08 | [-0.28, 0.43] |
| Timeafter | | -30.97 | [-54.72, -7.22] | -2.59 | 82 | .011* | -1.11 | [-1.96, -0.26] |
| Intervention reg | | -31.18 | [-356.59, 294.24] | -0.19 | 82 | .849 | -0.05 | [-0.55, 0.45] |
| Timeafter Intervention reg | | 9.24 | [0.01, 18.47] | 1.99 | 82 | .050* | 0.29 | [0.00, 0.59] |
| season [spring] | | -161.74 | [-332.72, 9.24] | -1.88 | 82 | .063 | -0.52 | [-1.06, 0.03] |
| season [summer] | | -192.69 | [-396.51, | -1.88 | 82 | .064 | -0.62 | [-1.27, |

| | | | | | | | | |
|---|---|---|---|---|---|---|---|---|
| | | | 11.12] | | | | | 0.04] |
| season [autumn] | | -330.47 | [-591.70, -69.24] | -2.52 | 82 | .014* | -1.06 | [-1.89, -0.22] |
| CPI change | | -36.64 | [-151.15, 77.87] | -0.64 | 82 | .526 | -0.31 | [-0.86, 0.25] |
| reg × CPI change | | -36.05 | [-151.85, 79.75] | -0.62 | 82 | .537 | -0.10 | [-0.43, 0.23] |
| AIC | 1,163.69 | | | | | | | |
| AICc | 1,168.24 | | | | | | | |
| BIC | 1,196.76 | | | | | | | |
| R2 | 0.89 | | | | | | | |
| R2 (adj.) | 0.88 | | | | | | | |
| Sigma | 110.05 | | | | | | | |

Authors' analysis of Kantar's Worldpanel OOH Purchase panel, 47w/e, 27th Nov 2022.

Autocorrelation:

```
## OK: Residuals appear to be independent and not autocorrelated (p = 0.360).
```

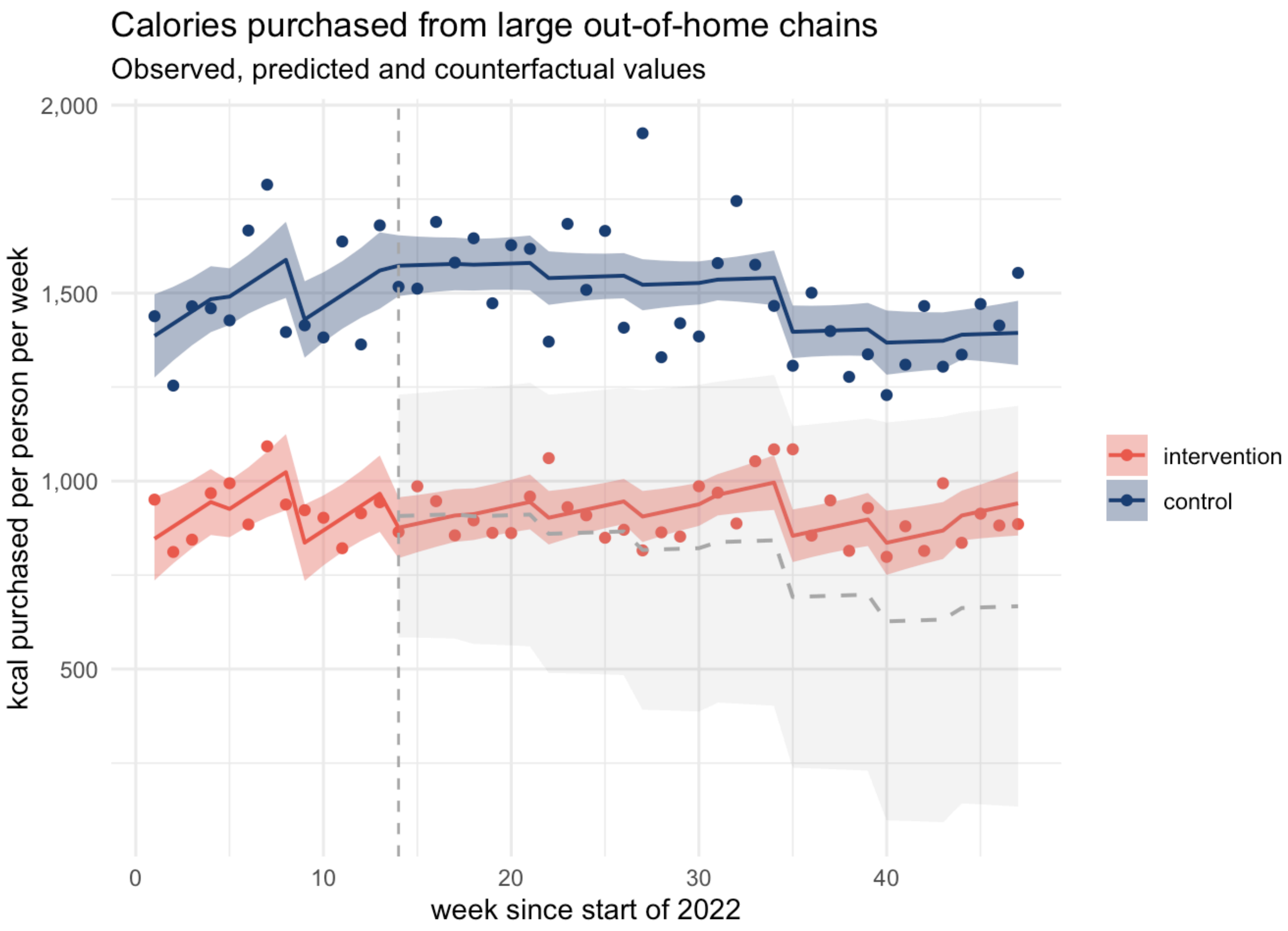


Population-level observed (points) and predicted (solid lines) calories per person purchased from large out-of-home (OOH) chains with counterfactual (grey dashed line) and 95% confidence intervals (ribbons). Implementation of mandatory calorie labelling = week 14. Large chains denote restaurant and takeaway businesses identified in the data that have ≥250 employees. The intervention series includes purchases made from large OOH chains in England. The control series is constructed from purchases made in Scotland and Wales and excludes purchases from large chains. Authors' analysis of Kantar's Worldpanel OOH Purchase panel, 47w/e, 27th Nov 2022.

### Purchases from non-chains

This outcome concerns purchases made from all OOH outlets excluding those that can be identified as large chain restaurants and takeaways required to show calories. These had been excluded already from the control series in all other models.

*Calories (kcal) per person per week purchased from non-chains*

| Parameter | Fit | *b* | 95% CI (*b*) | *t* | *df* | *p* | *b** | 95% CI (*b**) |
|---|---|---|---|---|---|---|---|---|
| (Intercept) | | 1,718.93 | [1076.98, 2360.88] | 5.33 | 82 | < .001*** | 1.44 | [0.23, 2.65] |
| reg | | -101.39 | [-815.03, 612.25] | -0.28 | 82 | .778 | -0.18 | [-1.34, 0.97] |
| Time | | 35.77 | [12.47, 59.07] | 3.05 | 82 | .003** | 3.65 | [1.27, 6.03] |
| Intervention | | 90.20 | [-151.38, 331.77] | 0.74 | 82 | .460 | 0.30 | [-0.51, 1.12] |
| Timeafter | | -31.02 | [-54.12, -7.92] | -2.67 | 82 | .009** | -2.60 | [-4.54, -0.66] |
| Intervention reg | | -154.98 | [-471.50, 161.54] | -0.97 | 82 | .333 | -0.56 | [-1.71, 0.58] |
| Timeafter Intervention reg | | 4.23 | [-4.75, 13.20] | 0.94 | 82 | .352 | 0.31 | [-0.35, 0.98] |
| season [spring] | | -133.14 | [-299.44, 33.17] | -1.59 | 82 | .115 | -1.00 | [-2.24, 0.25] |
| season [summer] | | -211.02 | [-409.26, -12.78] | -2.12 | 82 | .037* | -1.58 | [-3.06, -0.10] |

| | | | | | | | | |
|---|---|---|---|---|---|---|---|---|
| season [autumn] | | -349.20 | [-603.29, -95.11] | -2.73 | 82 | .008** | -2.61 | [-4.52, -0.71] |
| CPI change | | -62.31 | [-173.70, 49.07] | -1.11 | 82 | .269 | -0.78 | [-2.05, 0.48] |
| reg × CPI change | | 5.98 | [-106.66, 118.61] | 0.11 | 82 | .916 | 0.04 | [-0.71, 0.79] |
| AIC | 1,158.48 | | | | | | | |
| AICc | 1,163.03 | | | | | | | |
| BIC | 1,191.55 | | | | | | | |
| R2 | 0.43 | | | | | | | |
| R2 (adj.) | 0.36 | | | | | | | |
| Sigma | 107.04 | | | | | | | |

Authors' analysis of Kantar's Worldpanel OOH Purchase panel, 47w/e, 27th Nov 2022.

Autocorrelation:

```
## OK: Residuals appear to be independent and not autocorrelated (p = 0.238).
```

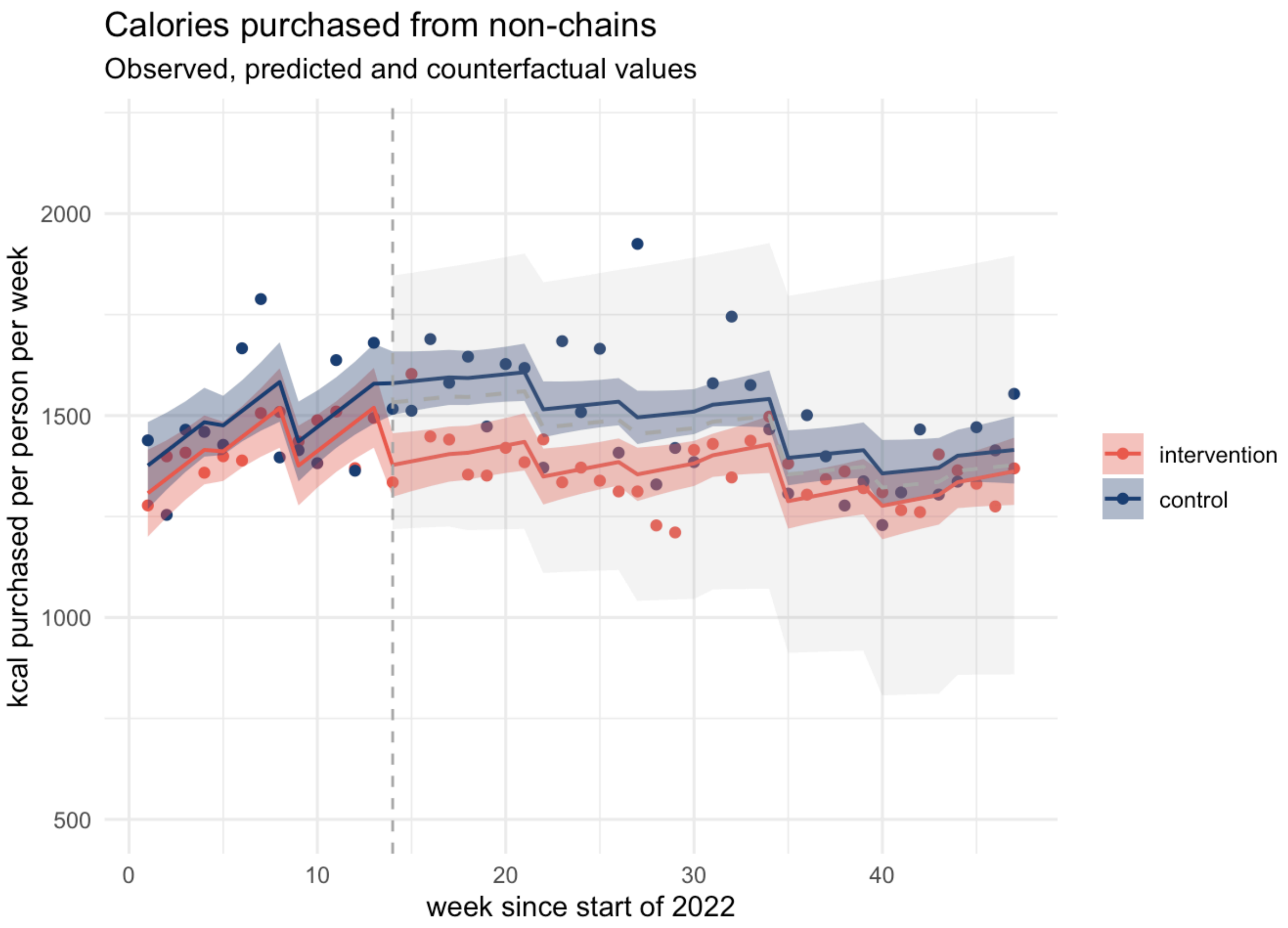


Population-level observed (points) and predicted (solid lines) calories purchased from out of home (OOH) food outlets other than large chains with counterfactual (grey dashed line) and 95% confidence intervals (ribbons). Implementation of mandatory calorie labelling = week 14. The intervention series includes all OOH purchases in England. The control series is constructed from purchases made in Scotland and Wales and excludes purchases from large chains. Large chains denote restaurant and takeaway businesses identified in the data that have ≥250 employees. Authors' analysis of Kantar's Worldpanel OOH Purchase panel, 47w/e, 27th Nov 2022.

### Meals

*Calories (kcal) per person per week from meals purchased out of home*

| Parameter | Fit | *b* | 95% CI (*b*) | *t* | *df* | *p* | *b** | 95% CI (*b**) |
|---|---|---|---|---|---|---|---|---|
| (Intercept) | | 1,308.33 | [667.18, 1949.48] | 4.06 | 82 | < .001*** | 0.52 | [-0.05, 1.09] |
| reg | | 660.81 | [-51.94, 1373.56] | 1.84 | 82 | .069 | 0.83 | [0.29, 1.38] |
| Time | | 33.49 | [10.21, 56.76] | 2.86 | 82 | .005** | 1.61 | [0.49, 2.73] |
| Intervention | | 81.19 | [-160.08, 322.46] | 0.67 | 82 | .505 | 0.13 | [-0.25, 0.51] |
| Timeafter | | -30.85 | [-53.93, -7.78] | -2.66 | 82 | .009** | -1.22 | [-2.13, -0.31] |
| Intervention reg | | -78.58 | [-394.70, 237.55] | -0.49 | 82 | .622 | -0.13 | [-0.67, 0.40] |
| Timeafter Intervention reg | | 8.27 | [-0.70, 17.23] | 1.83 | 82 | .070 | 0.29 | [-0.02, 0.60] |
| season [spring] | | -117.96 | [-284.06, 48.14] | -1.41 | 82 | .161 | -0.42 | [-1.00, 0.17] |
| season [summer] | | -155.96 | [-353.95, 42.04] | -1.57 | 82 | .121 | -0.55 | [-1.25, 0.15] |
| season [autumn] | | -263.31 | [-517.08, | -2.06 | 82 | .042* | -0.93 | [-1.82, |

| | | | | | | | | |
|---|---|---|---|---|---|---|---|---|
| | | | -9.53] | | | | | -0.03] |
| CPI change | | -70.23 | [-181.48, 41.01] | -1.26 | 82 | .213 | -0.50 | [-1.10, 0.09] |
| reg × CPI change | | -21.27 | [-133.77, 91.22] | -0.38 | 82 | .708 | -0.07 | [-0.42, 0.28] |
| AIC | 1,158.25 | | | | | | | |
| AICc | 1,162.80 | | | | | | | |
| BIC | 1,191.31 | | | | | | | |
| R2 | 0.88 | | | | | | | |
| R2 (adj.) | 0.86 | | | | | | | |
| Sigma | 106.91 | | | | | | | |

Authors’ analysis of Kantar’s Worldpanel OOH Purchase panel, 47w/e, 27th Nov 2022.

Autocorrelation:

```
## OK: Residuals appear to be independent and not autocorrelated (p = 0.878).
```

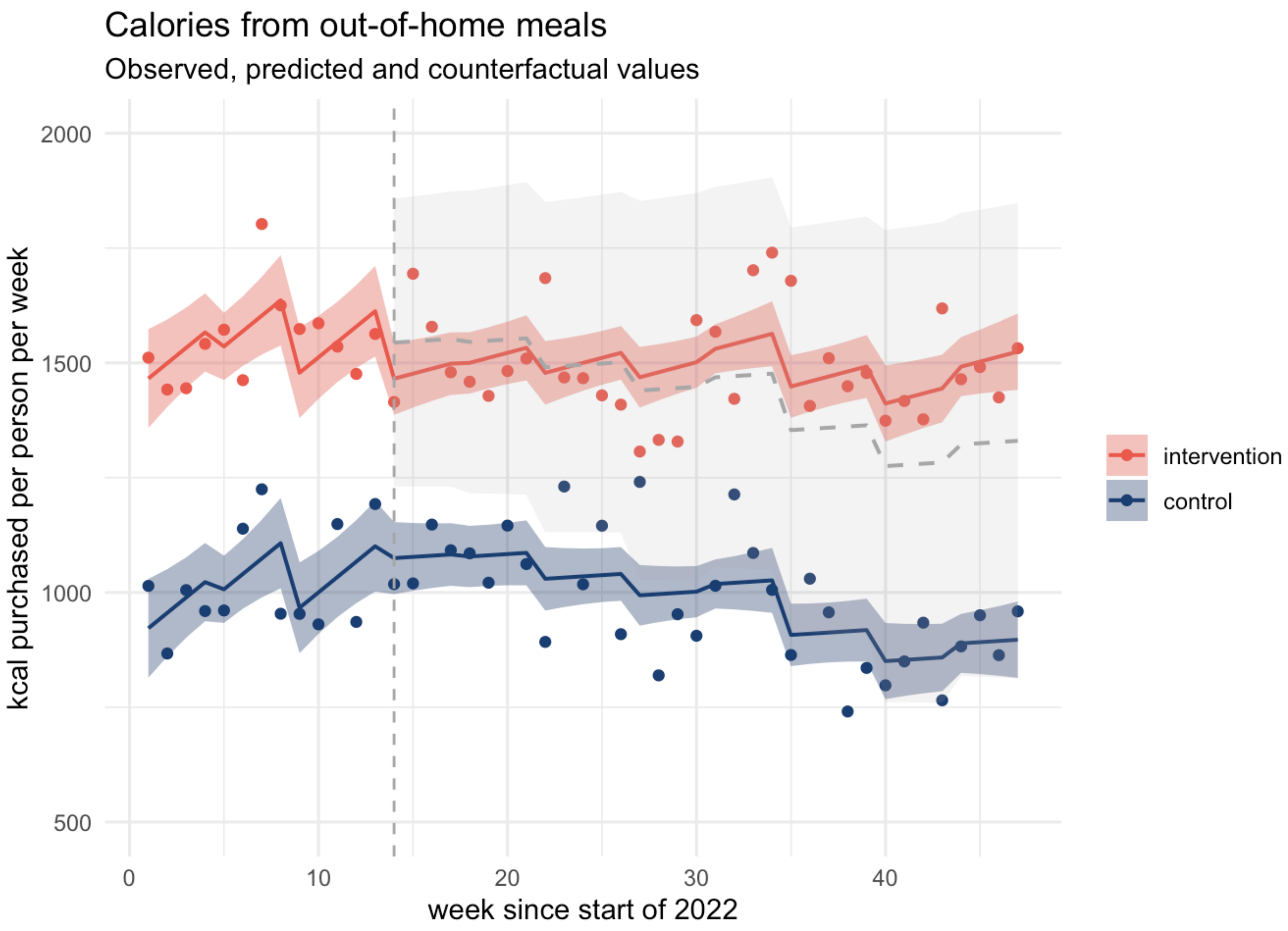


Population-level observed (points) and predicted (solid lines) calories purchased from out of home (OOH) meals with counterfactual (grey dashed line) and 95% confidence intervals (ribbons). Implementation of mandatory calorie labelling = week 14. The intervention series includes all OOH purchases in England. The control series is constructed from purchases made in Scotland and Wales and excludes purchases from large chains. Large chains denote restaurant and takeaway businesses identified in the data that have ≥250 employees. Authors' analysis of Kantar's Worldpanel OOH Purchase panel, 47w/e, 27th Nov 2022.

### Coffees

We distinguish between high-calorie coffees (high milk content, e.g. cappuccino, latte, moccha, frappe) and low-calorie coffees (filter coffee, espresso, macchiato).

*Calories (kcal) per person per week from higher-calorie coffee purchased out of home*

| Parameter | Fit | *b* | 95% CI (*b*) | *t* | *df* | *p* | *b** | 95% CI (*b**) |
|---|---|---|---|---|---|---|---|---|
| (Intercept) | | 84.59 | [40.07, 129.12] | 3.78 | 82 | < .001*** | 0.15 | [-0.39, 0.69] |
| reg | | 42.06 | [-7.44, 91.56] | 1.69 | 82 | .095 | 0.91 | [0.39, 1.43] |
| Time | | 0.99 | [-0.63, 2.61] | 1.22 | 82 | .226 | 0.65 | [-0.41, 1.72] |
| Intervention | | 7.56 | [-9.20, 24.31] | 0.90 | 82 | .372 | 0.16 | [-0.20, 0.53] |
| Timeafter | | -0.75 | [-2.35, 0.85] | -0.93 | 82 | .354 | -0.41 | [-1.27, 0.46] |
| Intervention reg | | -6.33 | [-28.28, 15.63] | -0.57 | 82 | .568 | -0.15 | [-0.66, 0.37] |
| Timeafter Intervention reg | | 0.44 | [-0.18, 1.06] | 1.40 | 82 | .165 | 0.21 | [-0.09, 0.51] |
| season [spring] | | -1.42 | [-12.96, 10.11] | -0.25 | 82 | .807 | -0.07 | [-0.63, 0.49] |
| season [summer] | | -0.76 | [-14.51, 12.99] | -0.11 | 82 | .912 | -0.04 | [-0.70, 0.63] |

| | | | | | | | | |
|---|---|---|---|---|---|---|---|---|
| season [autumn] | | -8.83 | [-26.46, 8.79] | -1.00 | 82 | .322 | -0.43 | [-1.28, 0.43] |
| CPI change | | -4.89 | [-12.62, 2.83] | -1.26 | 82 | .211 | -0.44 | [-1.01, 0.13] |
| reg × CPI change | | -0.52 | [-8.33, 7.29] | -0.13 | 82 | .895 | -0.02 | [-0.36, 0.31] |
| AIC | 656.82 | | | | | | | |
| AICc | 661.37 | | | | | | | |
| BIC | 689.89 | | | | | | | |
| R2 | 0.89 | | | | | | | |
| R2 (adj.) | 0.87 | | | | | | | |
| Sigma | 7.42 | | | | | | | |

Authors' analysis of Kantar's Worldpanel OOH Purchase panel, 47w/e, 27th Nov 2022.

Autocorrelation:

```
## OK: Residuals appear to be independent and not autocorrelated (p = 0.978).
```

*Calories (kcal) per person per week from lower-calorie coffee purchased out of home*

| Parameter | Fit | *b* | 95% CI (*b*) | *t* | *df* | *p* | *b** | 95% CI (*b**) |
|---|---|---|---|---|---|---|---|---|
| (Intercept) | | 4.36 | [-3.75, 12.46] | 1.07 | 82 | .288 | 0.99 | [-0.00, 1.98] |
| reg | | 8.59 | [-0.41, 17.60] | 1.90 | 82 | .061 | -1.54 | [-2.48, -0.59] |
| Time | | 0.29 | [-0.01, 0.58] | 1.93 | 82 | .057 | 1.89 | [-0.06, 3.84] |
| Intervention | | -3.22 | [-6.27, -0.17] | -2.10 | 82 | .039* | -0.70 | [-1.37, -0.04] |
| Timeafter | | -0.54 | [-0.83, -0.24] | -3.65 | 82 | < .001*** | -2.91 | [-4.49, -1.32] |
| Intervention reg | | 1.16 | [-2.84, 5.15] | 0.58 | 82 | .566 | 0.27 | [-0.67, 1.21] |
| Timeafter Intervention reg | | 0.30 | [0.18, 0.41] | 5.23 | 82 | < .001*** | 1.44 | [0.89, 1.98] |
| season [spring] | | -2.05 | [-4.15, 0.05] | -1.94 | 82 | .056 | -0.99 | [-2.01, 0.03] |
| season [summer] | | -2.70 | [-5.20, -0.19] | -2.14 | 82 | .035* | -1.31 | [-2.52, -0.09] |
| season [autumn] | | -2.63 | [-5.84, 0.58] | -1.63 | 82 | .107 | -1.28 | [-2.83, 0.28] |
| CPI change | | 1.58 | [0.17, 2.98] | 2.23 | 82 | .028* | 0.63 | [-0.40, |

| | | | | | | | | |
|---|---|---|---|---|---|---|---|---|
| | | | | | | | | 1.67] |
| reg × CPI change | | -1.67 | [-3.09, -0.25] | -2.34 | 82 | .022* | -0.72 | [-1.33, -0.11] |
| AIC | 336.51 | | | | | | | |
| AICc | 341.06 | | | | | | | |
| BIC | 369.57 | | | | | | | |
| R2 | 0.62 | | | | | | | |
| R2 (adj.) | 0.57 | | | | | | | |
| Sigma | 1.35 | | | | | | | |

Authors’ analysis of Kantar’s Worldpanel OOH Purchase panel, 47w/e, 27th Nov 2022.

Autocorrelation:

```
## OK: Residuals appear to be independent and not autocorrelated (p = 0.230).
```

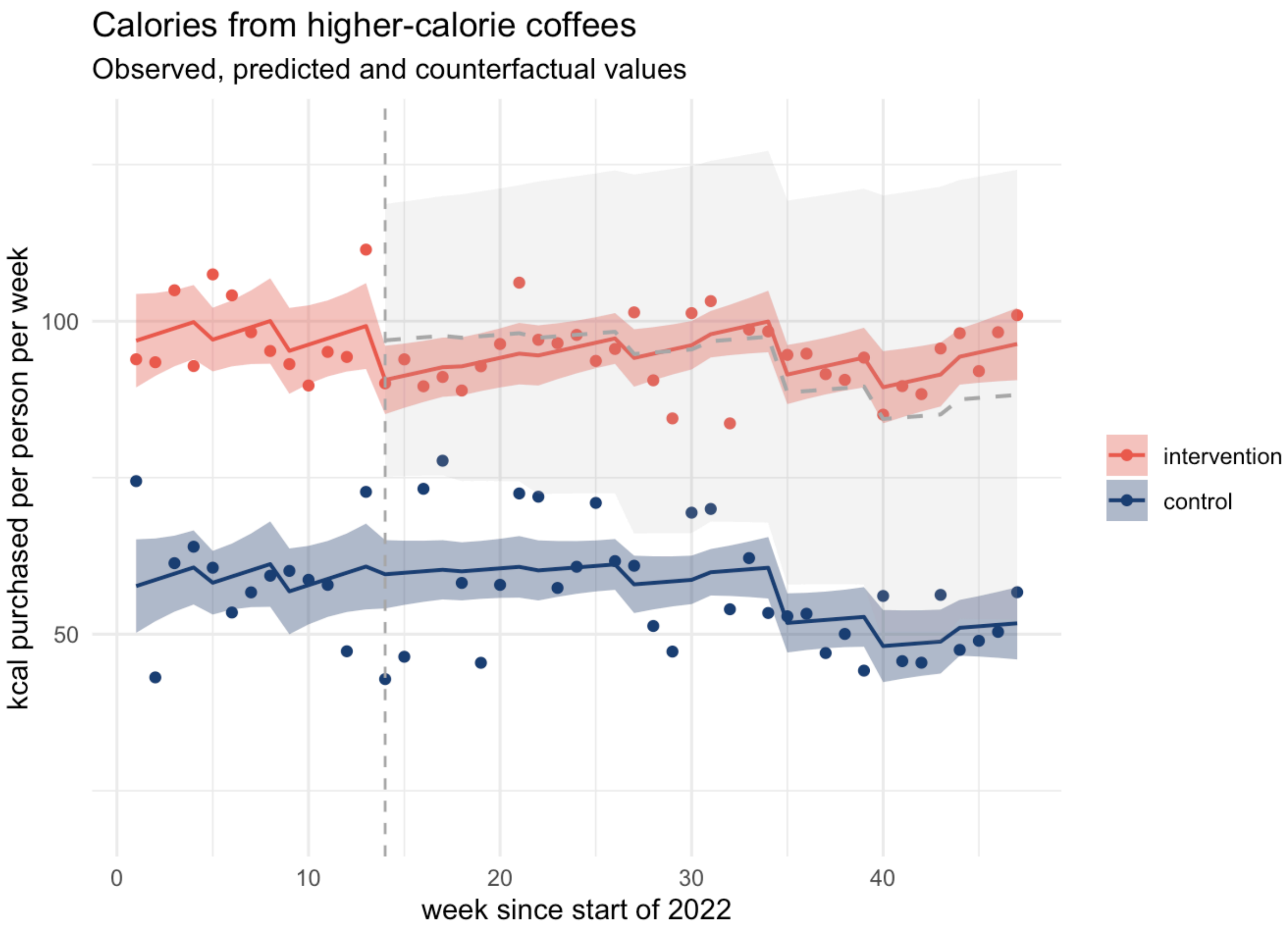


Population-level observed (points) and predicted (solid lines) calories per person purchased from higher-calorie coffees with counterfactual (grey dashed line) 95% confidence intervals (ribbons). Implementation of mandatory calorie labelling = week 14. The intervention series includes purchases in England. The control series is constructed from purchases made in Scotland and Wales and excludes purchases from large chains. Large chains denote restaurant and takeaway businesses identified in the data that have ≥250 employees. Authors' analysis of Kantar's Worldpanel OOH Purchase panel, 47w/e, 27th Nov 2022.

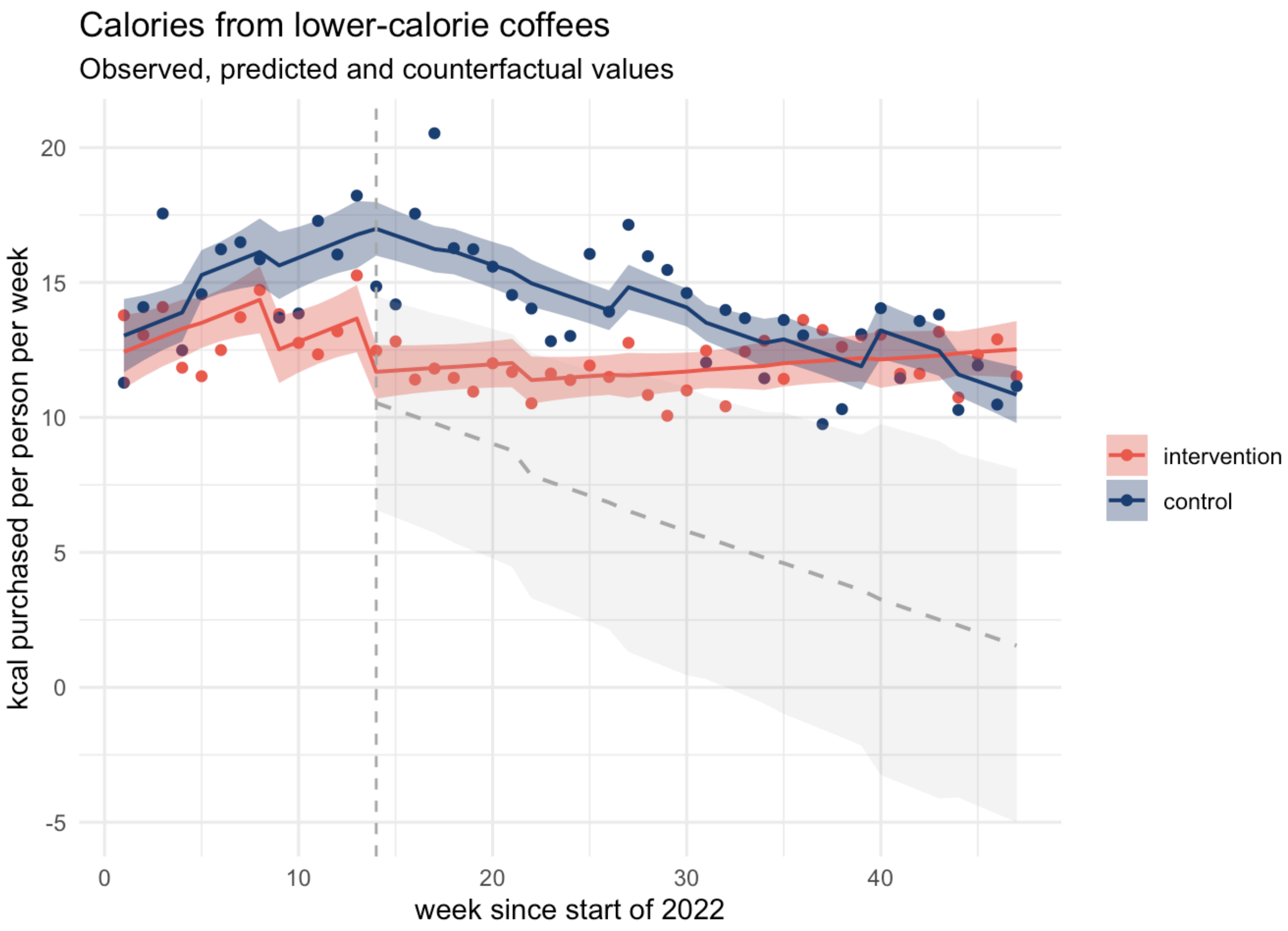


Population-level observed (points) and predicted (solid lines) calories per person purchased from lower-calorie coffees with counterfactual (grey dashed line) 95% confidence intervals (ribbons). Implementation of mandatory calorie labelling = week 14. The intervention series includes purchases in England. The control series is constructed from purchases made in Scotland and Wales and excludes purchases from large chains. Large chains denote restaurant and takeaway businesses identified in the data that have ≥250 employees. Authors' analysis of Kantar's Worldpanel OOH Purchase panel, 47w/e, 27th Nov 2022.

### Sandwiches

*Calories (kcal) per person per week from sandwiches purchased out of home*

| Parameter | Fit | *b* | 95% CI (*b*) | *t* | *df* | *p* | *b** | 95% CI (*b**) |
|---|---|---|---|---|---|---|---|---|
| (Intercept) | | 318.85 | [165.16, 472.53] | 4.13 | 82 | < .001*** | 0.30 | [-0.56, 1.17] |
| reg | | 15.64 | [-155.21, 186.49] | 0.18 | 82 | .856 | 1.31 | [0.48, 2.14] |
| Time | | 9.69 | [4.12, 15.27] | 3.46 | 82 | .001*** | 2.97 | [1.26, 4.68] |
| Intervention | | 25.88 | [-31.95, 83.71] | 0.89 | 82 | .376 | 0.26 | [-0.32, 0.84] |
| Timeafter | | -6.77 | [-12.30, -1.24] | -2.44 | 82 | .017* | -1.70 | [-3.09, -0.31] |
| Intervention reg | | -31.70 | [-107.48, 44.08] | -0.83 | 82 | .408 | -0.34 | [-1.17, 0.48] |
| Timeafter Intervention reg | | -1.95 | [-4.10, 0.19] | -1.81 | 82 | .074 | -0.44 | [-0.92, 0.04] |
| season [spring] | | -11.99 | [-51.80, 27.83] | -0.60 | 82 | .551 | -0.27 | [-1.16, 0.62] |
| season [summer] | | -16.06 | [-63.52, 31.40] | -0.67 | 82 | .503 | -0.36 | [-1.43, 0.70] |
| season [autumn] | | -20.96 | [-81.79, 39.87] | -0.69 | 82 | .495 | -0.47 | [-1.84, |

| | | | | | | | | |
|---|---|---|---|---|---|---|---|---|
| | | | | | | | | 0.89] |
| CPI change | | -33.85 | [-60.51, -7.18] | -2.52 | 82 | .013* | -1.12 | [-2.02, -0.21] |
| reg × CPI change | | 11.24 | [-15.72, 38.21] | 0.83 | 82 | .409 | 0.22 | [-0.31, 0.76] |
| AIC | 889.72 | | | | | | | |
| AICc | 894.27 | | | | | | | |
| BIC | 922.78 | | | | | | | |
| R2 | 0.71 | | | | | | | |
| R2 (adj.) | 0.67 | | | | | | | |
| Sigma | 25.63 | | | | | | | |

Authors' analysis of Kantar's Worldpanel OOH Purchase panel, 47w/e, 27th Nov 2022.

Autocorrelation:

```
## OK: Residuals appear to be independent and not autocorrelated (p = 0.092).
```

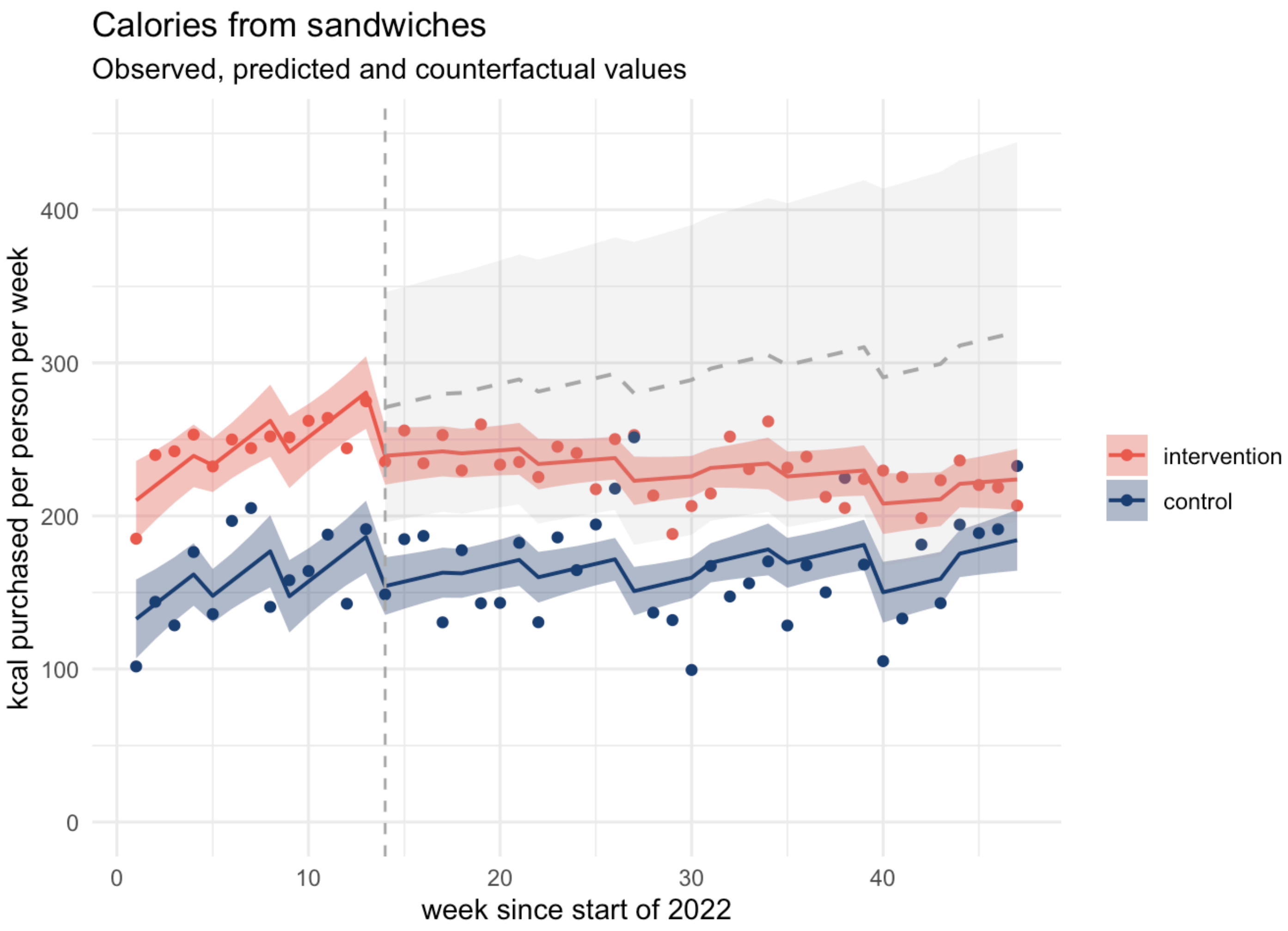


Population-level observed (points) and predicted (solid lines) calories per person purchased from sandwiches with counterfactual (grey dashed line) 95% confidence intervals (ribbons). Implementation of mandatory calorie labelling = week 14. The intervention series includes purchases in England. The control series is constructed from purchases made in Scotland and Wales and excludes purchases from large chains. Large chains denote restaurant and takeaway businesses identified in the data that have ≥250 employees. Authors' analysis of Kantar's Worldpanel OOH Purchase panel, 47w/e, 27th Nov 2022.

### Fish and chip meals

*Calories (kcal) per person per week from fish and chip meals purchased out of home*

| Parameter | Fit | *b* | 95% CI (*b*) | *t* | *df* | *p* | *b** | 95% CI (*b**) |
|---|---|---|---|---|---|---|---|---|
| (Intercept) | | 127.94 | [-8.89, 264.77] | 1.86 | 82 | .066 | 0.38 | [-0.99, 1.75] |
| reg | | 21.76 | [-130.35, 173.87] | 0.28 | 82 | .777 | 0.06 | [-1.24, 1.37] |
| Time | | 1.13 | [-3.84, 6.09] | 0.45 | 82 | .653 | 0.61 | [-2.08, 3.30] |
| Intervention | | -21.81 | [-73.30, 29.68] | -0.84 | 82 | .402 | -0.39 | [-1.31, 0.53] |
| Timeafter | | -1.16 | [-6.08, 3.77] | -0.47 | 82 | .642 | -0.51 | [-2.70, 1.68] |
| Intervention reg | | 13.22 | [-54.25, 80.68] | 0.39 | 82 | .698 | 0.25 | [-1.04, 1.55] |
| Timeafter Intervention reg | | 0.13 | [-1.78, 2.04] | 0.13 | 82 | .893 | 0.05 | [-0.70, 0.81] |
| season [spring] | | -6.27 | [-41.72, 29.18] | -0.35 | 82 | .726 | -0.25 | [-1.66, 1.16] |
| season [summer] | | -5.54 | [-47.80, 36.71] | -0.26 | 82 | .795 | -0.22 | [-1.90, 1.46] |
| season [autumn] | | -22.91 | [-77.07, 31.24] | -0.84 | 82 | .402 | -0.91 | [-3.06, |

| | | | | | | | | |
|---|---|---|---|---|---|---|---|---|
| | | | | | | | | 1.24] |
| CPI change | | 0.56 | [-23.18, 24.30] | 0.05 | 82 | .963 | -0.03 | [-1.47, 1.40] |
| reg × CPI change | | -2.09 | [-26.10, 21.92] | -0.17 | 82 | .863 | -0.07 | [-0.92, 0.77] |
| AIC | 867.88 | | | | | | | |
| AICc | 872.43 | | | | | | | |
| BIC | 900.94 | | | | | | | |
| R2 | 0.28 | | | | | | | |
| R2 (adj.) | 0.18 | | | | | | | |
| Sigma | 22.82 | | | | | | | |

Authors’ analysis of Kantar’s Worldpanel OOH Purchase panel, 47w/e, 27th Nov 2022.

Autocorrelation:

```
## OK: Residuals appear to be independent and not autocorrelated (p = 0.806).
```

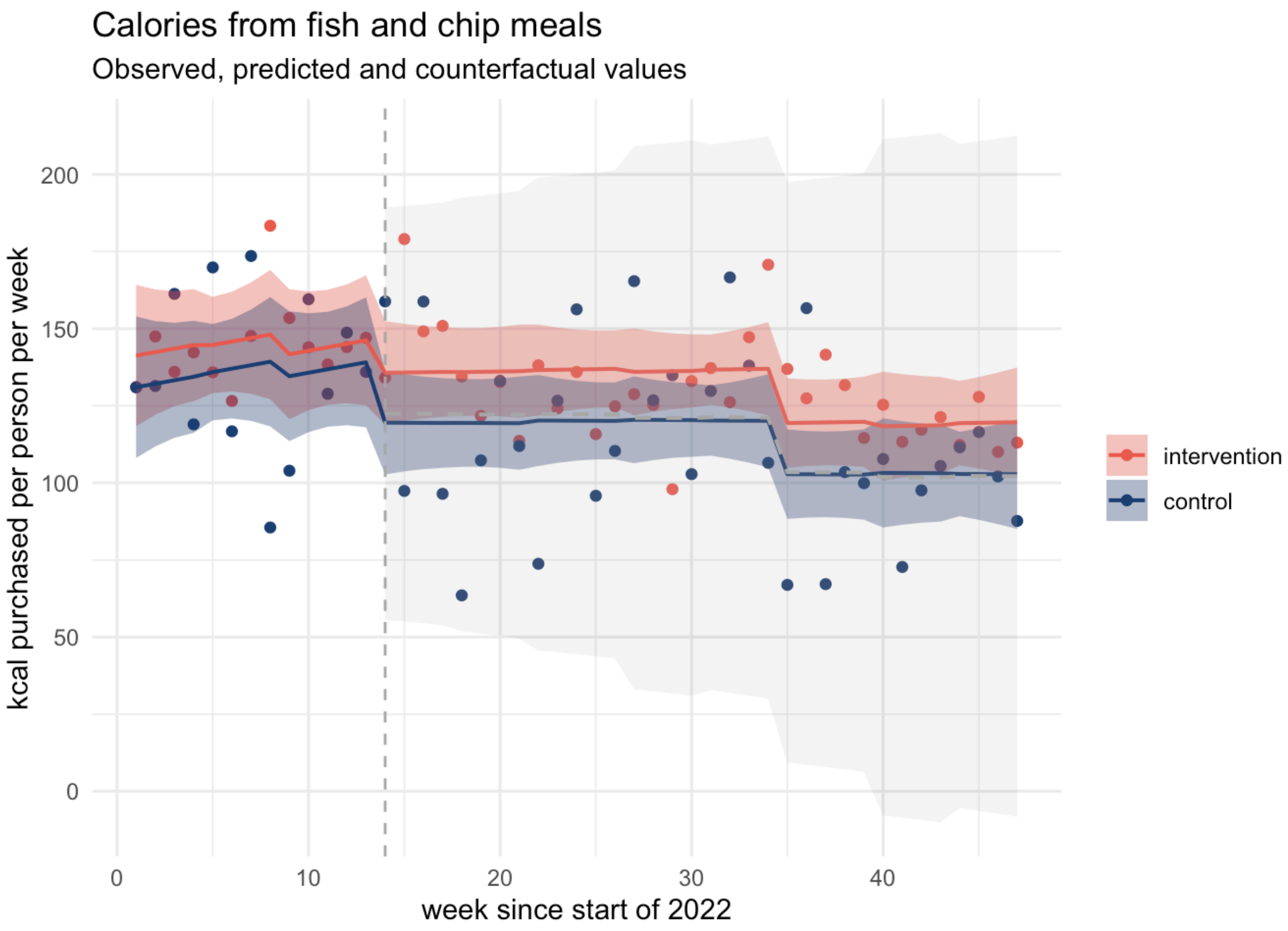


Population-level observed (points) and predicted (solid lines) calories per person purchased from fish and chip meals with counterfactual (grey dashed line) 95% confidence intervals (ribbons). Implementation of mandatory calorie labelling = week 14. The intervention series includes purchases in England. The control series is constructed from purchases made in Scotland and Wales and excludes purchases from large chains. Large chains denote restaurant and takeaway businesses identified in the data that have ≥250 employees. Authors' analysis of Kantar's Worldpanel OOH Purchase panel, 47w/e, 27th Nov 2022.
