## Supplementary Material 3 for "Changes in out-of-home food purchasing following England’s calorie labelling regulations: a population-level controlled interrupted time series"

### Description of calories purchased out of home by subgroup

**Table S2**. Unadjusted kcal purchased OOH per person and week by subgroups

| Subgroup | Average kcal purchased from OOH venues per person per week (SD) | | | | | | | |
|---|---|---|---|---|---|---|---|---|
| | Intervention series | | | | Control series | | | |
| Age | # underlying reporters | Pre intervention | Post intervention | Δ pre- and post-intervention | # underlying reporters | Pre intervention | Post intervention | Δ pre- and post-intervention |
| < 35 years | 1,071 | 2,239.3 (219.2) | 2,008.37 (186.3) | -230.9* | 95 | 865.5 (320.9) | 856.0 (244.9) | -9.4 |
| 35–54 years | 2,754 | 2,335.3 (187.2) | 2,296.6 (109.2) | -38.7 | 334 | 1,255.9 (230.6) | 1,361.1 (188.0) | 105.2 |
| 55+ years | 1,962 | 2,456.5 (113.8) | 2,493.1 (132.1) | 36.6 | 292 | 1.964.8 (141.6) | 1,949.0 (204.1) | -15.7 |
| Sex | | Pre intervention | Post intervention | | | Pre intervention | Post intervention | |
| Women | 3,617 | 2,300.8 (137.7) | 2,265.0 (151.7) | -35.9 | 436 | 1,481.7 (240.4) | 1,457.3 (182.5) | -24.4 |
| Men | 2,170 | 2,396.9 (110.1) | 2.282.0 (145.3) | -114.9* | 285 | 1,498.3 (139.1) | 1,519.0 (232.1) | 20.7 |
| SES | | Pre intervention | Post intervention | | | Pre intervention | Post intervention | |
| High | 3,745 | 2,381.2 (116.7) | 2,278.9 (165.5) | -102.3* | 450 | 1,516.7 (192.4) | 1,561.4 (166.3) | 44.8 |
| Low | 2,039 | 2,292.3 (153.9) | 2,265.0 (147.4) | -27.3 | 270 | 1,456.7 (218.0) | 1,396.4 (244.5) | -60.4 |
| Weight status | | | | | | | | |
| Under and healthy weight | 1,691 | 2,415.2 (173.4) | 2,255.1 (157.6) | -160.1* | 184 | 1,355.2 (162.9) | 1,354.1 (240.6) | -1.1 |
| Overweight | 1,413 | 2,153.9 (143.2) | 2,180.8 (168.6) | 26.9 | 192 | 1,433.0 (142.2) | 1,445.1 (229.7) | 12.1 |
| Obesity | 1,193 | 2,786.3 (187.8) | 2,696.5 (221.4) | -89.7 | 171 | 1,661.6 (327.3) | 1,688.7 (293.6) | 27.1 |
| | Average kcal purchased from OOH venues per person per day (SD) | | | | | | | |
| Day of the week | | Pre intervention | Post intervention | | | Pre intervention | Post intervention | |
| Weekday | 5,113 | 351.3 (27.9) | 346.5 (26.8) | -4.8 | 609 | 234.3 (28.1) | 230.9 (28.1) | -3.4 |
| Weekend | 5,219 | 865.4 (59.1) | 814.8 (43.5) | -50.5* | 636 | 526.9 (62.7) | 519.0 (74.6) | -7.9 |

OOH = out of home; SES = socio-economic status. The intervention series includes all out of home (OOH) purchases in England. The control series is constructed from purchases made in Scotland and Wales and excludes purchases from large chains. Large chains denote restaurant and takeaway businesses identified in the data that have ≥250 employees. Authors' analysis of Kantar's Worldpanel OOH Purchase panel, 47w/e, 27th Nov 2022. OOH = out-of-home. * $p<0.05$ (two-sample t-test, unequal variances).
