## Supplementary Material 4 for "Changes in out-of-home food purchasing following England’s calorie labelling regulations: a population-level controlled interrupted time series"

### Robustness checks

This document contains results from the robustness checks of 'Changes in out-of-home food purchasing following England's calorie labelling regulations: a population-level controlled interrupted time series'.

#### 1 Including varying pre-intervention trends

In this robustness check, we included an interaction term between pre-intervention time and group (intervention/control), thereby letting time trends vary by group.

**Table S3**. Immediate (level) and longer-term (slope) effects of mandatory calorie labelling at population level – sensitivity analysis 1: including varying pre-intervention trends

| Outcome (kcal per person per week) | Level change (95%CI) | P | Slope change (95%CI) | P |
|---|---|---|---|---|
| All calories purchased OOH | -67.8 (-477.01 to 341.5) | 0.743 | -1.3 (-35.9 to 33.4) | 0.943 |
| From large chains [a] | -80.5 (-428.8 to 267.8) | 0.647 | 20.6 (-8.9 to 50.1) | 0.169 |
| From non-chains | -163.8 (-498.8 to 171.2) | 0.338 | 6.3 (-22.1 to 34.6) | 0.666 |
| From meals | -76.2 (-415.9 to 263.5) | 0.656 | 7.7 (-21.0 to 36.5) | 0.595 |
| From higher-calorie coffees | -3.8 (-27.4 to 19.7) | 0.747 | -0.14 (-2.1 to 1.7) | 0.892 |
| From lower-calorie coffees | 1.5 (-2.8 to 5.8) | 0.481 | 0.21 (-0.15 to 0.58) | 0.246 |
| From sandwiches | -47.3 (-128.2 to 33.5) | 0.247 | 1.6 (-5.2 to 8.5) | 0.637 |
| From fish and chip meals | 27.6 (-44.4 to 99.5) | 0.448 | -3.2 (-9.3 to 2.9) | 0.305 |

95%CI = 95% confidence interval; OOH = out of home. [a] Large chains denote restaurant and takeaway businesses identified in the data that have ≥250 employees. Authors' analysis of Kantar's Worldpanel OOH Purchase panel, 47w/e, 27th Nov 2022.

#### 2 Purchases made for the individual themselves only

This robustness check considers only purchases reported by the individual as purchased for themselves only (57.8% of all transactions). The main analysis also includes purchases made by the individual for other adults and children.

**Table S4**. Immediate (level) and longer-term (slope) effects of mandatory calorie labelling at population level – sensitivity analysis 2: purchases made for the individual themselves only

| Outcome (kcal per person per week) | Level change (95%CI) | P | Slope change (95%CI) | P |
|---|---|---|---|---|
| All calories purchased OOH | -105.8 (-278.6 to 66.9) | 0.230 | -1.2 (-6.1 to 3.8) | 0.647 |
| From large chains [a] | -51.5 (-210.4 to 107.4) | 0.525 | 2.0 (-2.5 to 6.5) | 0.387 |
| From non-chains | -116.2 (-279.6 to 47.3) | 0.164 | -1.3 (-6.0 to 3.3) | 0.575 |
| From meals | -70.5 (-201.9 to 61.0) | 0.294 | **3.7 (0.01 to 7.5)** | **0.050** |
| From higher-calorie coffees | -4.5 (-19.2 to 10.1) | 0.544 | **0.47 (0.06 to 0.89)** | **0.026** |
| From lower-calorie coffees | 1.9 (-1.5 to 5.3) | 0.279 | **0.27 (0.17 to 0.36)** | **<0.001** |
| From sandwiches [b] | -22.4 (-77.1 to 32.3) | 0.423 | **-2.5 (-4.3 to -0.64)** | **0.008** |
| From fish and chip meals | 1.9 (-36.7 to 40.4) | 0.925 | -0.19 (-1.3 to 0.92) | 0.740 |

95%CI = 95% confidence interval; OOH = out of home. [a] Large chains denote restaurant and takeaway businesses identified in the data that have ≥250 employees. [b] Autocorrelation detected and robust standard errors used. Authors' analysis of Kantar's Worldpanel OOH Purchase panel, 47w/e, 27th Nov 2022.

#### 3 Excluding top 1% & 10% high-calorie purchase occasions

In this robustness check we removed purchases made during the purchase occasions with the top 1% / top 10% highest calorie count of items purchased during the occasion. This allower-calories us to assess the impact of these outlier purchases on the estimated intervention effect.

**Table S5**. Immediate (level) and longer-term (slope) effects of mandatory calorie labelling at population level – sensitivity analysis 3a: excluding top 1% of high-calorie purchase occasions

| Outcome (kcal per person per week) | Level change (95%CI) | P | Slope change (95%CI) | P |
|---|---|---|---|---|
| All calories purchased OOH | -110.8 (-409.2 to 187.5) | 0.467 | 1.5 (-7.0 to 9.8) | 0.734 |
| From large chains | -49.0 (-318.8 to 220.8) | 0.722 | 5.4 (-2.2 to 13.1) | 0.165 |
| From non-chains [a] | -143.7 (-408.7 to 121.2) | 0.288 | 1.0 (-6.5 to 8.5) | 0.792 |
| From meals | -91.2 (-327.4 to 144.9) | 0.449 | 4.8 (-2.0 to 11.5) | 0.165 |
| From higher-calorie coffees | -5.7 (-27.2 to 15.8) | 0.602 | 0.47 (-0.14 to 1.1) | 0.130 |
| From lower-calorie coffees | 1.1 (-2.8 to 5.0) | 0.592 | **0.30 (0.19 to 0.41)** | **<0.001** |
| From sandwiches | -30.7 (-105.6 to 44.2) | 0.422 | -1.9 (-4.0 to 0.22) | 0.078 |
| From fish and chip meals | -15.4 (-66.2 to 35.5) | 0.554 | -0.7 (-2.2 to 0.7) | 0.329 |

95%CI = 95% confidence interval; OOH = out of home. [a] Large chains denote restaurant and takeaway businesses identified in the data that have ≥250 employees. Authors' analysis of Kantar's Worldpanel OOH Purchase panel, 47w/e, 27th Nov 2022.

**Table S6**. Immediate (level) and longer-term (slope) effects of mandatory calorie labelling at population level – sensitivity analysis 3b: excluding top 10% of high-calorie purchase occasions

| Outcome (kcal per person per week) | Level change (95%CI) | P | Slope change (95%CI) | P |
|---|---|---|---|---|
| All calories purchased OOH | -119.1 (-272.3 to 34.1) | 0.128 | -2.8 (-7.1 to 1.6) | 0.212 |
| From large chains [a] | -91.1 (-228.6 to 46.5) | 0.194 | 0.11 (-3.7 to 3.9) | 0.956 |
| From non-chains | -118.8 (-263.0 to 25.4) | 0.106 | -2.9 (-6.9 to 1.2) | 0.173 |
| From meals | **-97.0 (-188.8 to -5.2)** | **0.038** | 0.8 (-1.8 to 3.4) | 0.560 |
| From higher-calorie coffees | -5.7 (-26.1 to 14.6) | 0.580 | 0.47 (-0.10 to 1.1) | 0.107 |
| From lower-calorie coffees | 2.4 (-1.1 to 5.8) | 0.178 | **0.30 (0.21 to 0.40)** | **<0.001** |
| From sandwiches | -28.8 (-102.4 to 44.8) | 0.443 | -2.0 (-4.1 to 0.05) | 0.055 |
| From fish and chip meals | -4.8 (-27.3 to 17.7) | 0.673 | **-1.3 (-2.0 to -0.57)** | **<0.001** |

95%CI = 95% confidence interval; OOH = out of home. [a] Large chains denote restaurant and takeaway businesses identified in the data that have ≥250 employees. Authors' analysis of Kantar's Worldpanel OOH Purchase panel, 47w/e, 27th Nov 2022.

#### 4 Balanced observations

In this robustness check, we restrict the post-intervention series to 13 weeks to match the pre-intervention series.

**Table S7**. Immediate (level) and longer-term (slope) effects of mandatory calorie labelling at population level – sensitivity analysis 4: using balanced observations pre and post labelling

| Outcome (kcal per person per week) | Level change (95%CI) | P | Slope change (95%CI) | P |
|---|---|---|---|---|
| All calories purchased OOH [a] | -123.1 (-490.9 to 244.7) | 0.512 | -8.1 (-34.9 to 18.6) | 0.552 |
| From large chains [b] | 11.2 (-367.0 to 389.3) | 0.954 | 4.9 (-17.3 to 27.0) | 0.667 |
| From non-chains | -107.5 (-478.4 to 263.5) | 0.570 | -7.1 (-28.9 to 14.6) | 0.519 |
| From meals | -61.0 (-421.5 to 299.6) | 0.740 | -4.6 (-25.7 to 16.5) | 0.669 |
| From higher-calorie coffees | -5.7 (-36.4 to 25.1) | 0.718 | -0.4 (-2.2 to 1.4) | 0.645 |
| From lower-calorie coffees | -0.49 (-5.7 to 4.7) | 0.855 | 0.21 (-0.09 to 0.52) | 0.175 |
| From sandwiches | 4.7 (-73.8 to 83.3) | 0.906 | -3.4 (-8.0 to 1.2) | 0.147 |
| From fish and chip meals | 10.1 (-76.4 to 96.5) | 0.820 | -1.9 (-7.0 to 3.1) | 0.459 |

95%CI = 95% confidence interval; OOH = out of home. [a] Autocorrelation detected and robust standard errors used. [b] Large chains denote restaurant and takeaway businesses identified in the data that have ≥250 employees. Authors' analysis of Kantar's Worldpanel OOH Purchase panel, 47w/e, 27th Nov 2022.

#### 5 Temporal falsification

In this robustness check, we moved the intervention 'backwards' by four weeks to the week commencing 7th March 2022.

**Table S8**. Immediate (level) and longer-term (slope) effects of mandatory calorie labelling at population level – sensitivity analysis 5: temporal falsification (intervention 4 weeks earlier)

| Outcome (kcal per person per week) | Level change (95%CI) | P | Slope change (95%CI) | P |
|---|---|---|---|---|
| All calories purchased OOH | 0.24 (-267.5 to 268.0) | 0.999 | 6.9 (-4.1 to 17.8) | 0.218 |
| From large chains [a] | -70.4 (-299.1 to 158.3) | 0.547 | 8.7 (-0.63 to 18.0) | 0.068 |
| From non-chains | 29.9 (-192.8 to 252.6) | 0.793 | 7.4 (-1.7 to 16.5) | 0.109 |
| From meals | -58.8 (-280.8 to 163.2) | 0.604 | 8.6 (-0.49 to 17.6) | 0.064 |
| From higher-calorie coffees | 0.54 (-14.8 to 15.9) | 0.945 | 0.59 (-0.03 to 1.2) | 0.063 |
| From lower-calorie coffees | -0.19 (-3.1 to 2.7) | 0.895 | **0.29 (0.18 to 0.41)** | **<0.001** |
| From sandwiches | -3.9 (-57.0 to 49.1) | 0.884 | -1.7 (-3.8 to 0.49) | 0.128 |
| From fish and chip meals | -17.2 (-63.9 to 29.5) | 0.470 | -0.43 (-2.3 to 1.5) | 0.655 |

95%CI = 95% confidence interval; OOH = out of home. [a] Large chains denote restaurant and takeaway businesses identified in the data that have ≥250 employees. Authors' analysis of Kantar's Worldpanel OOH Purchase panel, 47w/e, 27th Nov 2022.

#### 6 Including price index

This robustness check includes indices for prices of food and drink items purchased out of home created from the purchase data. This index was expressed as average spend per 100 kcal across all items purchased in each week and region, and does not account for sales.

**Table S9**. Immediate (level) and longer-term (slope) effects of mandatory calorie labelling at population level – sensitivity analysis 6: including a price index of average price per item

| Outcome (kcal per person per week) | Level change (95%CI) | P | Slope change (95%CI) | P |
|---|---|---|---|---|
| All calories purchased OOH | -76.3 (-453.4 to 300.7) | 0.692 | 6.26 (-4.6 to 17.1) | 0.258 |
| From large chains [a] | -13.7 (-335.2 to 307.8) | 0.934 | **10.3 (1.0 to 19.5)** | **0.030** |
| From non-chains | -135.4 (-447.4 to 176.5) | 0.395 | 5.4 (-3.6 to 14.3) | 0.242 |
| From meals | -70.5 (-384.8 to 243.7) | 0.660 | 8.7 (-0.3 to 17.8) | 0.058 |
| From higher-calorie coffees | -7.6 (-29.2 to 14.1) | 0.495 | 0.37 (-0.26 to 0.99) | 0.247 |
| From lower-calorie coffees | 1.2 (-2.8 to 5.2) | 0.556 | **0.30 (0.19 to 0.41)** | **<0.001** |
| From sandwiches | -22.9 (-95.6 to 49.8) | 0.537 | -1.5 (-3.5 to 0.65) | 0.175 |
| From fish and chip meals | 12.1 (-55.1 to 79.2) | 0.725 | 0.06 (-1.9 to 2.0) | 0.949 |

95%CI = 95% confidence interval; OOH = out of home. [a] Large chains denote restaurant and takeaway businesses identified in the data that have ≥250 employees. Authors' analysis of Kantar's Worldpanel OOH Purchase panel, 47w/e, 27th Nov 2022.
